# Dominant negative ATP5F1A variants disrupt oxidative phosphorylation causing neurological disorders

**DOI:** 10.1101/2025.07.08.25330848

**Authors:** Sara M. Fielder, Marisa W. Friederich, Daniella H. Hock, Jessie R. Zhang, Liana M. Valin, Jill A. Rosenfeld, Kevin T.A. Booth, Natasha J. Brown, Rocio Rius, Tanavi Sharma, Liana N. Semcesen, Kim C. Worley, Lindsay C. Burrage, Kayla Treat, Tara Samson, Sarah Govert, Sara DaCunha, Weimin Yuan, Jian Chen, Jacob Lesinski, Hieu Hoang, Stephanie A. Morrison, Farah A. Ladha, Roxanne A. Van Hove, Cole R. Michel, Richard Reisdorph, Eric Tycksen, Dustin Baldridge, Gary A. Silverman, Claudia Soler-Alfonso, Erin Conboy, Francesco Vetrini, Lisa Emrick, William J. Craigen, Undiagnosed Diseases Network, Stephen M. Sykes, David A. Stroud, Johan L.K. Van Hove, Tim Schedl, Stephen C. Pak

**Author notes:** Co-senior author (Johan L.K. Van Hove, Tim Schedl, Stephen C. Pak).

## Abstract

*ATP5F1A* encodes the α-subunit of complex V of the respiratory chain, which is responsible for mitochondrial ATP synthesis. We describe 6 probands with heterozygous *de novo* missense *ATP5F1A* variants that presented with developmental delay, intellectual disability, and movement disorders. Functional evaluation in *C. elegans* revealed that all variants tested were damaging to gene function via a dominant negative genetic mechanism. Biochemical and proteomics studies showed a marked reduction in complex V abundance and activity in proband-derived blood cells and fibroblasts. Mitochondrial physiology studies in fibroblasts revealed increased oxygen consumption, yet decreased mitochondrial membrane potential and ATP levels indicative of uncoupled oxidative phosphorylation as a pathophysiologic mechanism. Our findings contrast functionally and clinically with the previously reported *ATP5F1A* variant, p.Arg207His, suggesting a distinct pathological mechanism. This study therefore expands the phenotypic and genotypic spectrum of *ATP5F1A*-associated conditions and highlights how functional studies can provide understanding of the genetic, molecular, and cellular mechanisms of *ATP5F1A* variants of uncertain significance. With 12 heterozygous individuals now reported, *ATP5F1A* is the most frequent nuclear genome cause of complex V deficiency.

## INTRODUCTION

Adenosine triphosphate (ATP) generation and utilization is a highly coordinated and tightly regulated process. Central to this process is mitochondrial ATP synthase, also known as complex V, which is an essential enzyme in cellular metabolism that is responsible for producing ∼90% of cellular ATP. ATP synthase is comprised of a central domain that contains a membrane embedded central ring (F_o_) organized by repeating c subunits, attached via a central stalk consisting of the δ, γ and ε subunits to a central globular head (F_1_) (Fig. 1) (Dautant et al., 2018; Galber et al., 2021). Three α- and three β-subunits, encoded by *ATP5F1A* and *ATP5F1B*, respectively, come together to form a hetero-hexameric ring which has the catalytic activity domain, where ATP synthesis occurs. A peripheral arm consisting of the b, d, and F6 subunits links to the head through the ATP5PO (also known as oligomycin sensitivity-conferring protein (OSCP)) subunit, and is membrane embedded with the a, e, f, g, j, k, and 8 subunits. The latter contains the proton pore. The enzyme operates via chemiosmosis which couples the proton gradient generated by the electron transport chain (ETC) with ATP synthesis. Movement of protons from the intermembrane space to the mitochondrial matrix drives the rotation of the central stalk (Boyer, 1997; Kuhlbrandt, 2019; Yoshida et al., 2001). Rotation of the central stalk then induces conformational changes in the α and β subunits of the F_1_ complex allowing coordinated binding of ADP and inorganic phosphate (Pi) and the catalysis of ATP. One complete rotation of the central stalk produces 3 molecules of ATP (Boyer, 1997).

**Figure 1.**
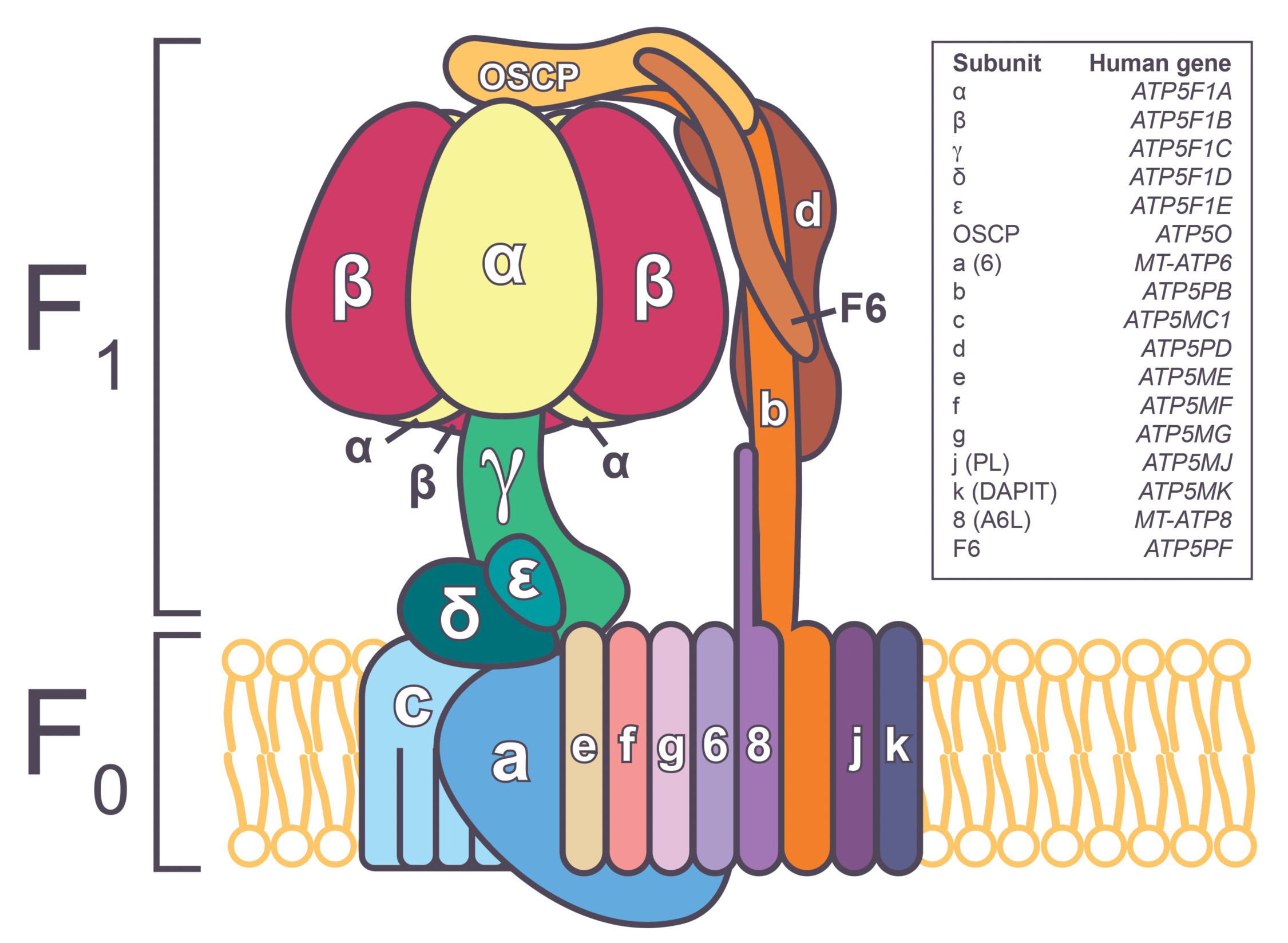
A schematic of ATP synthase (Complex V). Human mitochondrial ATP synthase is comprised of two domains, F_1_ and F_o_. F_1_ consists of α, β, γ, δ, and ε-subunits at a ratio of 3:3:1:1:1, and forms the catalytic activity domain where ATP synthesis occurs. F_o_ forms the proton pore and consists of a membrane embedded central ring organized by repeating c subunits, and a, e, f, g, j, k, and 8 subunits. A peripheral arm consisting of the b, d, and F6 subunits links to the catalytic head through the ATP5PO (also known as oligomycin sensitivity-conferring protein (OSCP)) subunit, and is embedded in the inner mitochondrial membrane.

Genetic variants in *ATP5F1A* associated with human disease have rarely been reported. Two families with biallelic pathogenic variants p.Tyr321Cys and p.Arg329Cys in *ATP5F1A* presented with neonatal or early infantile fatal conditions of devastating neurological lesions, pulmonary hypertension, and cardiac failure (Jonckheere et al., 2013; Lieber et al., 2013). Fibroblasts homozygous for the latter variant showed reduced complex V hydrolytic activity, with decreased amounts of the α, β, and ATP5PO subunits (Lieber et al., 2013). The parents of these children were unaffected. In contrast, two reports described a recurrent *de novo* heterozygous missense variant p.Arg207His in *ATP5F1A* with life-threatening neonatal-onset mitochondriopathy with failure to thrive, hyperlactatemia, hyperammonemia with elevated glutamine, and decreased citrulline (Lines et al., 2021; Zech et al., 2022). Fibroblasts derived from these individuals displayed reduced ATP5F1A protein, reduced complex V hydrolytic enzyme activity, a lower oxygen consumption rate, and normal inner mitochondrial membrane potential. All three affected individuals showed apparent clinical resolution by 18 months of age with normal development, although the long-term prognosis is currently unknown. In a second publication, the same p.Arg207His variant was described in a child with transient developmental delay, failure to thrive and lactic acidemia, and reduced complex V activity. Two other dominant *de novo* variants in *ATP5F1A* p.Arg182Gln and p.Ser346Phe were also reported in individuals who presented with developmental delay, cognitive dysfunction, and dystonia (Zech et al., 2022). Functional studies of these variants were not performed.

Here we describe six probands with heterozygous *de novo* missense variants in *ATP5F1A*, representing four variants, p.Arg182Gln, p.Ser346Phe, p.Pro331Leu, and p.Leu109Ser. All six probands presented with complex but overlapping neurological phenotypes including developmental delays, intellectual disability, pyramidal tract dysfunction, and dystonia. *In vivo* functional studies in *C. elegans* provided strong evidence that these variants are damaging to function by a dominant negative mechanism. Furthermore, through protein modeling, proteomics analysis, biochemical and mitochondrial physiology studies using proband-derived cells, we provide new insights into their molecular and cellular mechanism in fibroblasts which are distinct from the previously published dominant (p.Arg207His) and recessive (p.Tyr321Cys and p.Arg329Cys) *ATP5F1A* variants. Our study expands the genetic, clinical, and biochemical description of recurring nuclear complex V defects associated with pathogenic *ATP5F1A* variants.

## RESULTS

### Proband presentations

Proband 1 is a male with dystonia, global developmental delay since birth, and mild to moderate intellectual disability (Table 1). He was diagnosed with cerebral palsy and walks with the aid of braces. His first words were delayed. His examination is significant for dystonia, spasticity, gait apraxia, fluency disorder, poor fine and gross motor skills, limited attention, poor muscle tone, and brisk reflexes with normal sensation. He is nondysmorphic. Blood lactic acid levels were mildly elevated. Clinical duo exome sequencing with the father followed by targeted maternal studies identified a heterozygous *de novo* missense variant in *ATP5F1A* (NM_004046.6:c.545G>A p.(Arg182Gln)).

**Table 1.**
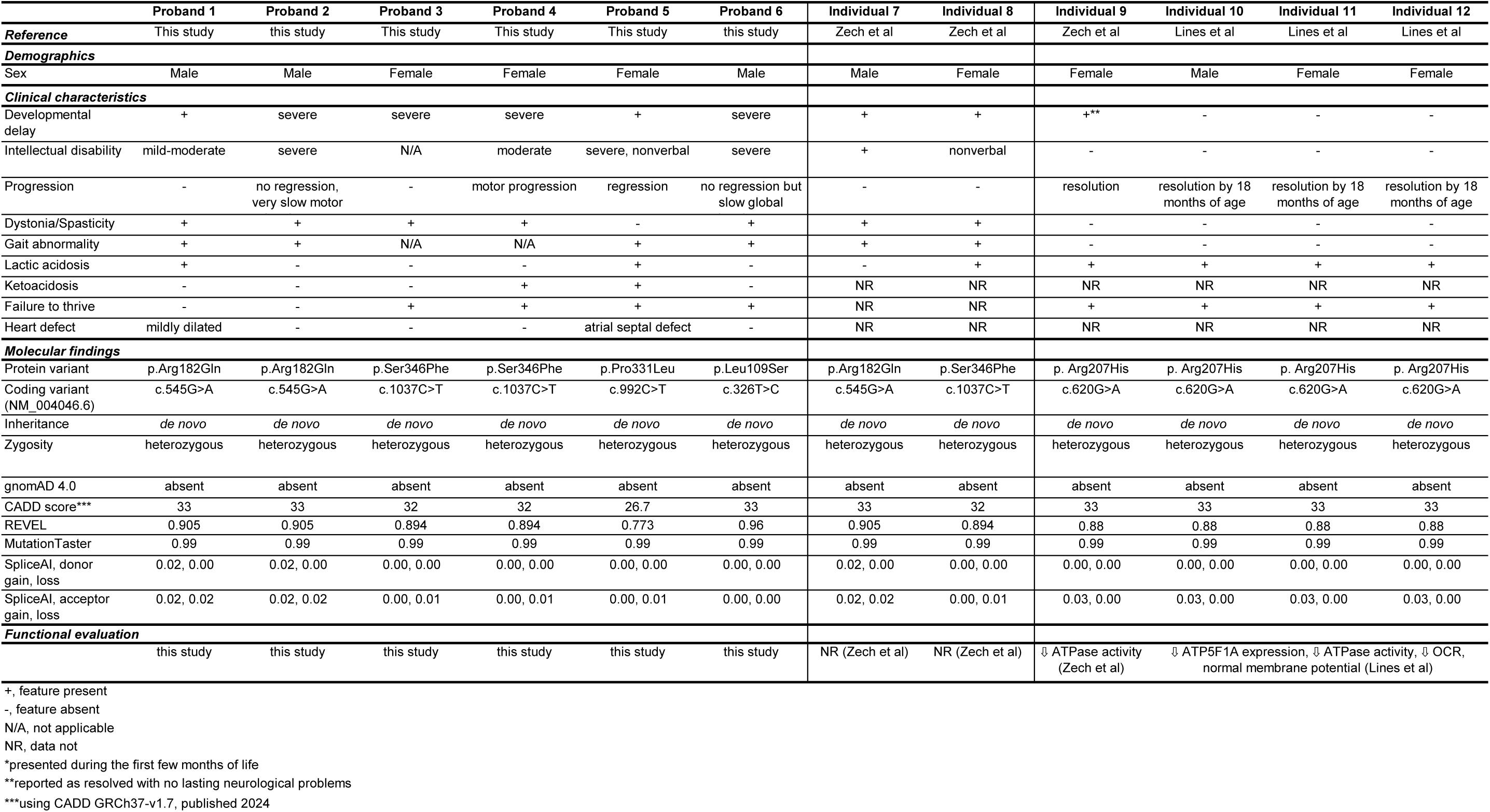
Clinical characteristics and molecular findings in individuals with ATP5F1A variants.

Proband 2 is a male with moderate to severe global developmental delay. Early history included significant central hypotonia with delayed crawling. He can walk with support, but independent ambulation has not been achieved. He has reduced tone with brisk reflexes in upper and lower limbs and upgoing plantar responses, tented upper lip with hypotonic facies and profound central hypotonia. Verbal communication is limited to a handful of words; however, receptive language appears better than expressive language. There has been no regression nor seizures. Growth is in the normal range. Trio genome sequencing identified a *de novo* heterozygous missense variant in *ATP5F1A* (NM_004046.6:c.545G>A p.(Arg182Gln)).

Proband 3 is a female with a significant history of severe global developmental delay and abnormal muscle tone, characterized by central hypotonia and peripheral spasticity. Her medical history is further complicated by dysphagia, chronic constipation, frequent vomiting, and feeding difficulties necessitating the placement of a gastrostomy tube (G-tube). She had a mildly elevated lactate, which at other times was normal. Clinical exome sequencing identified a heterozygous variant of uncertain significance in *ATP5F1A* (NM_004046.6:c.1037C>T p.(Ser346Phe)). Subsequent parental testing confirmed that the *ATP5F1A* variant in this patient is *de novo*.

Proband 4 is a female born prematurely. Developmental delays were noted in the first year of life. Over the next years, she gradually developed spasticity and dystonia, and could only walk with adaptive devices. On exam, she has dystonia most notable in facial and upper extremity movements. She has severe spasticity, most pronounced in her lower extremities, and has contractures in elbows and knees. She is fed by gastrostomy tube for dysphagia and has severe gastroesophageal reflux. While non-fasting, she had elevated ketones with 3-hydroxybutyrate 1.96 mM and acetoacetate 0.50 mM, and an elevated ratio 3.89 (normal 0.47-2.57). At age 11-15, she has growth retardation with height at -4 SD and weight at -3.3 SD. Genetic testing identified a *de novo* heterozygous variant in *ATP5F1A* (NM_004046.6:c.1037C>T p.(Ser346Phe)).

Proband 5 is a female with significant intellectual disability, developmental regression, growth failure, and history of hospitalizations for ketoacidosis. Prenatally, she was noted to have a dilated right ventricle in her brain, thought to be due to an ischemic event. Neonatally, she had hyperbilirubinemia, which improved with phototherapy. She had failure to thrive, an atrial septal defect, sinus tachycardia with normal QTc, and mild scoliosis. Developmental concerns were raised in the first year of life. She walked with stiffness and toe walking, which has improved. She is currently nonverbal. She has had 8 hospitalizations with ketoacidosis (3-hydroxybutyrate 3.6 mM), typically with normal ammonia, with the first episode requiring PICU admission. Her ketones normalize when she is healthy. She also has intermittent elevations of blood lactate (1.1-5.5 mM) with normal lactate:pyruvate ratio and urine lactate, without a clinical correlate. Clinical trio exome sequencing identified a *de novo* heterozygous missense variant in *ATP5F1A* (NM_004046.6:c.992C>T p.(Pro331Leu)).

Proband 6 is male with global developmental delay, limited verbal communication, postnatal failure to thrive with feeding difficulties, axial hypotonia, and leg spasticity with contractures (and therefore is nonambulatory). Trio whole genome sequencing identified a *de novo* heterozygous missense variant in *ATP5F1A* (NM_004046.6:c.326T>C p.(Leu109Ser)).

All six probands reported here show a shared phenotype of failure to thrive, developmental delay, and a motor syndrome of pyramidal tract involvement and dystonia. Lactic acid has been variable and mildly elevated, but two probands had excessive ketosis. A more comprehensive clinical report for each proband is available in the Supplementary Information.

The phenotypes observed in our six probands are in clear contrast to the phenotypes of the previously reported recurrent p.Arg207His variant, suggesting phenotypic expansion. Moreover, all individuals with p.Arg207His variant had neonatal onset mitochondriopathy that showed clinical resolution in late infancy (Lines et al., 2021; Zech et al., 2022), whereas the six probands in the current study have phenotypes that are either static or worsening (Table 1).

### Bioinformatic features of *ATP5F1A* variants

Clinical exome or whole genome sequencing identified *de novo* heterozygous missense variants in *ATP5F1A* (NM_004046.6) as the most likely candidate disease gene for each of the six cases (Table 1). The variants are absent from gnomAD 4.1 and predicted to be deleterious by *in silico* analysis: (CADD, all >25), pathogenic (REVEL, all >0.7), and disease causing (MutationTaster), while SpliceAI did not predict any effect on splicing (Table 1) (Ioannidis et al., 2016; Jaganathan et al., 2019; Schwarz et al., 2014; Zech et al., 2022). *ATP5F1A* is expected to be tolerant to loss-of-function variants (pLI=0.29; o/e=0.3 – 0.58), thus it is unlikely that haploinsufficiency is a genetic mechanism for *ATP5F1A*-related disease. Conversely, gnomAD data suggests that *ATP5F1A* is likely intolerant to missense variation (Z=3.16; o/e=0.63-0.73) (Chen et al., 2024).

### Structural analysis: proband variants are located at the contact points between α:β and α:γ subunits of the F1 complex of ATP synthase

To better understand the mechanism by which the proband variants might impact ATP synthase function, we mapped the locations of the variants to the cryoelectron microscopy-derived structure of the protein (Lai et al., 2023). All four affected residues, L109, R182, P331, and S346 are located at or near the contact points between α:β (ATP5F1A:ATP5F1B) or α:γ (ATP5F1A:ATP5F1C) subunits of the F1 complex (Fig. 2A and B). The P331 residue lies adjacent to P330 and is located in a pocket near the contact point with the β- and γ-subunits (Fig. 2C). Prolines are often found in turns and loops, and their distinctive cyclic sidechains provide conformational rigidity. While no hydrogen bonds are created or destroyed, the substitution of P331 with leucine dislocates the residue away from the β-subunit and moves it closer to the γ-subunit (Fig. 2D). This displacement, along with a change in flexibility is likely to compromise the structure of this region and alter interactions between the α-, β- and/or γ-subunits.

**Figure 2.**
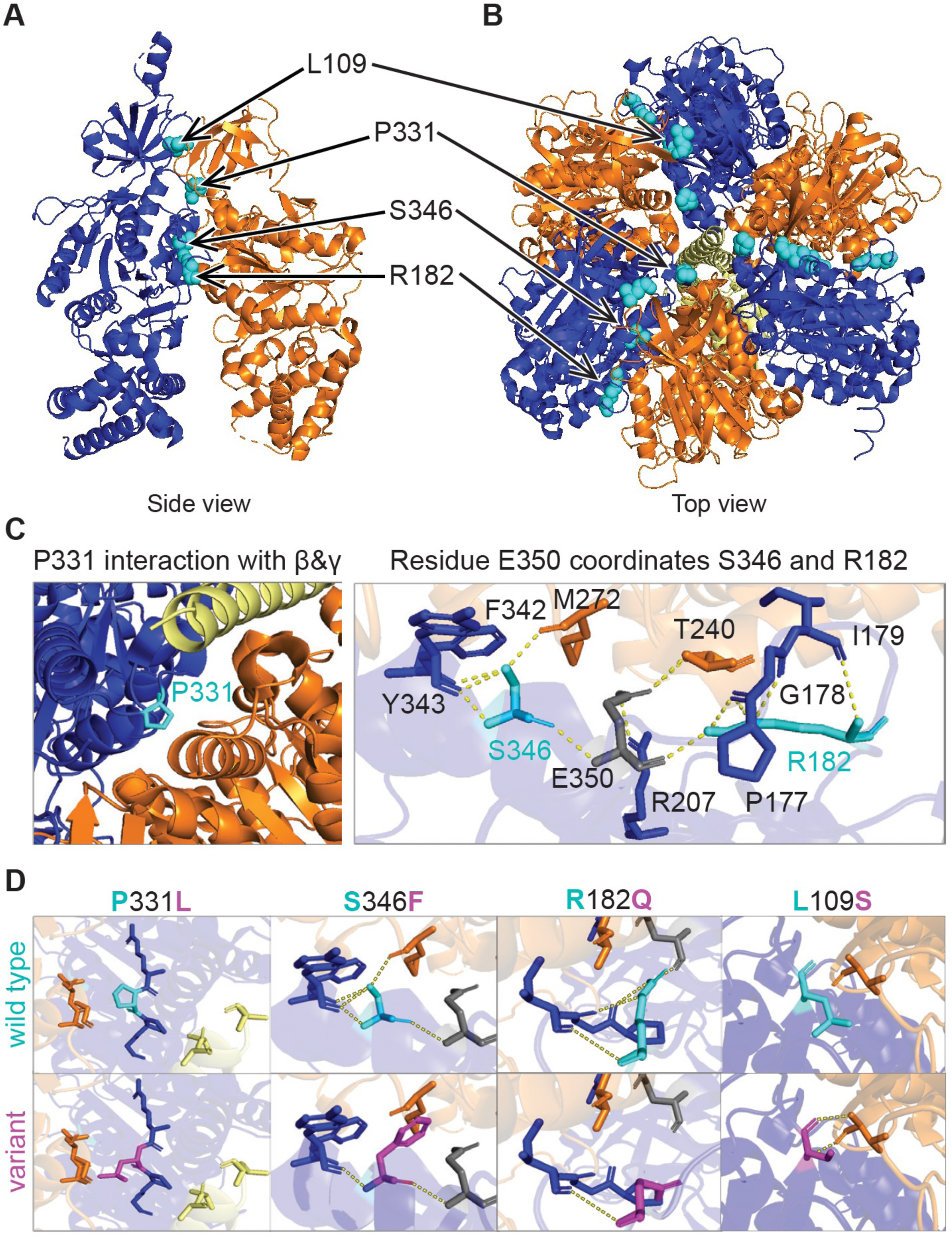
Proband *ATP5F1A* variants likely disrupt interaction between α, β, and γ subunits. Proband variants are located at the interfaces of α:β, and α:γ subunits. Locations of the four variants in a single α-β dimer (A), or in the hexameric complex with the γ subunit (B). α-subunit is depicted in dark blue, β-subunit in orange, γ-subunit in yellow, and variant locations are shown in cyan. (C) Close up schematic of wild type and variant interactions in the holocomplex. Wild type residues in cyan, proband variant residues in magenta, other residues in α-, β- and γ-subunits are in dark blue, orange, and yellow, respectively. The P331L, S346F and R182Q substitutions are predicted to destroy H-bonds and/or impact the conformation and interaction between α, β, and γ subunits. The L109S substitution creates two novel H-bonds between α, and β subunits which likely alters the interaction between these subunits.

The S346 residue interacts directly with the β-subunit via a hydrogen bond with M272 (β-subunit). S346 also forms multiple intra-α-subunit hydrogen bonds with E350, F342, and Y343 residues (Fig. 2C). The S346F substitution leads to the loss of hydrogen bonds with Y343 (α-subunit) and M272 (β-subunit) (Fig. 2D). M272 (β-subunit) lies adjacent to the hydrophobic pocket (aa 274-28) of the β-subunit, which is predicted to be critical for the interaction with the γ-subunit (Abrahams et al., 1994; Ganetzky et al., 2022). Therefore, the loss of the hydrogen bond with M272 (β-subunit) likely alters interaction between all three subunits.

Within the α-subunit, the R182 residue forms multiple hydrogen bonds with residues: E350, P177, G178, and I179 (Fig. 2C and D). The R182Q substitution is predicted to result in loss of hydrogen bonds with E350, P177, and G178 (Fig. 2D). The loss of three intra-α-subunit hydrogen bonds will likely have a significant impact on the protein confirmation. Interestingly, both S346 and R182 form hydrogen bonds with a common partner E350 (α-subunit), which interacts directly with T240 on the β-subunit via a hydrogen bond (Fig. 2C). This interaction with a common partner, E350, suggests that the S346F and R182Q variants act in a similar mechanism by impacting interaction with the β-subunit. The wild-type L109 residue is located at the interface between α- and β-subunits. The substitution of the non-polar, hydrophobic leucine with a polar, hydrophilic serine results in the formation of two new hydrogen bonds between the mutant S109 and V26 in the β-subunit. These changes likely alter the conformation of this region and change the interaction between these subunits (Fig. 2D). We hypothesize that these structural changes caused by the proband substitutions impact ATP synthase function by 1) disrupting proper formation of the ATP synthase complex, 2) impairing ADP + P_i_ binding, ATP synthesis or release, and/or 3) disrupting interaction between the γ-stalk with α- and β-subunits within the complex and thereby uncoupling the proton motive force from ATP synthesis. The possible disruption of complex V subunit interactions suggests that the *ATP5F1A* variants may have a change of function genetic mechanism.

### Proband variants are damaging to ATP-1 function *in vivo*

To determine if the proband variants are damaging *in vivo* and uncover the genetic mechanism of dominance observed in the probands, we modeled three variants (p.Arg182Gln, p.Pro331Leu, and p.Ser346Phe) in *C. elegans*. The fourth proband variant, p.Leu109Ser, was identified late in the project and thus was not modeled. *ATP5F1A* is highly conserved as *atp-1* in *C. elegans*, with 78% identity and 88% similarity between human and *C. elegans* proteins (Fig. S1). All three variant residues are conserved in *C. elegans* in regions of high homology (Fig. 3A and S1). This degree of conservation enabled us to employ CRISPR-Cas9 genome editing to knock in the proband variants into the corresponding *atp-1* residues (Arg167, Pro316, and Ser331) and evaluate the functional consequences of the missense variants *in vivo* (Fig. 3B).

**Figure 3.**
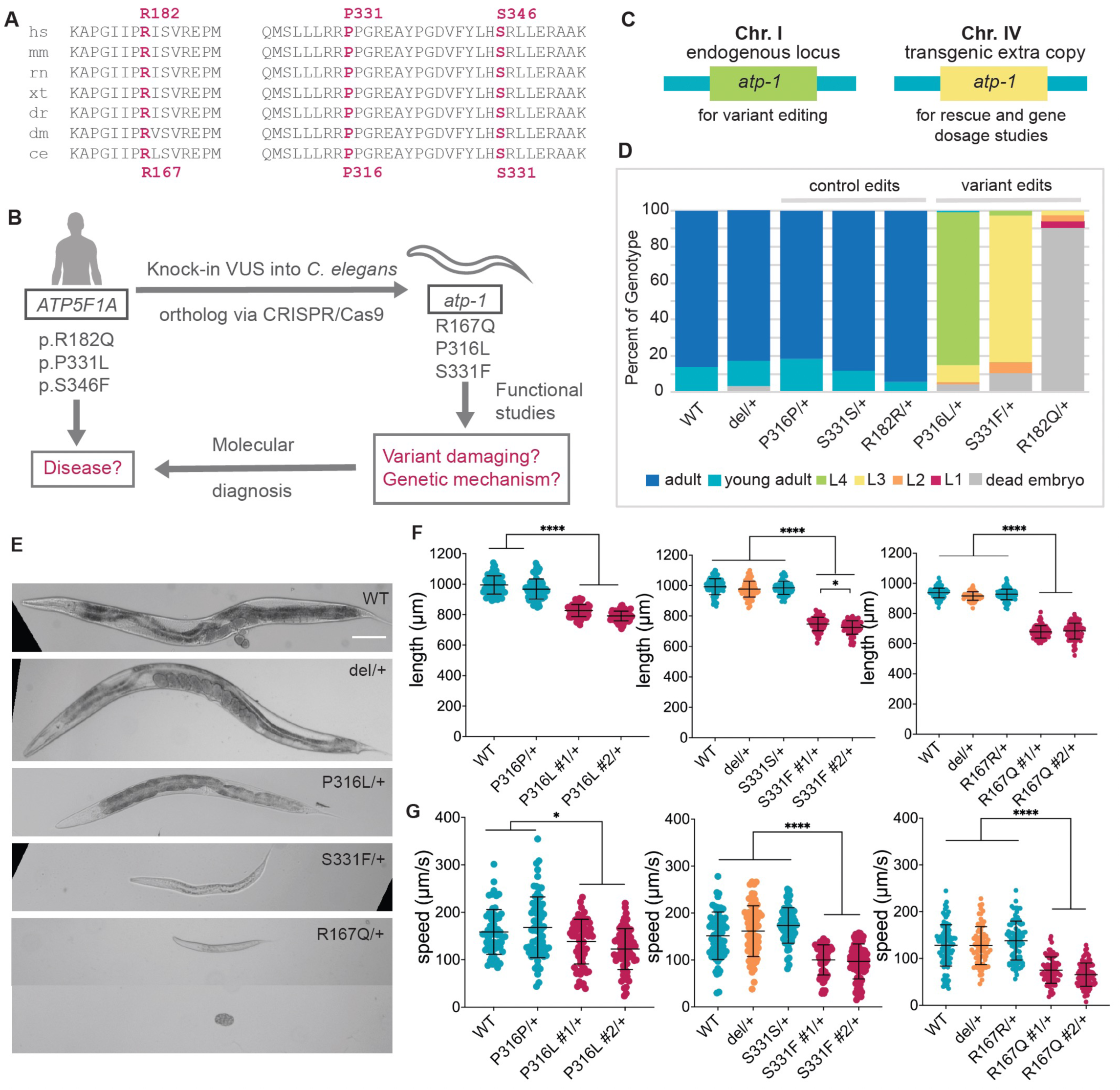
**P316L, S331F, and R167Q variants are damaging to *atp-1* function in *C. elegans.*** (A) R182, P331, and S346 residues are in areas of high conservation across multiple species, and all three residues are conserved in the *C. elegans* homolog *atp-1*. hs- human, mm- mouse, rn- rat, xt- xenopus, dr- zebrafish, dm- drosophila, ce- *C. elegans.* (B) Schematic of how variant modeling in *C. elegans* aid in molecular diagnosis of rare disease. (C) Schematic of *atp-1* extra copy lines generated that have endogenous *atp-1* on chromosome I, and a single copy transgene insertion of wild type *atp-1* on chromosome IV. (D) Development of heterozygous variant animals scored at 72 hours after embryo laying. Average of 4 experiments graphed, 60 embryos were counted in each replicate, except for R167Q, where at least 35 embryos were used. More information on the experiment and the crossing scheme used to generate these animals can be found in the Materials and Methods and in Fig. S3. (E) Representative images of animals in D. Scale bar, 100µm. (F) Length of heterozygous variant animals 24 hours post L4 larval stage as measured by WormLab over a two-minute recording. (G) Crawling speed of heterozygous variant animals 24 hours post L4 larval stage from the same recording as F. * p <0.05, ** p<0.01, *** p<0.001, **** p < 0.0001.

*atp-1* deletion (null) homozygotes were first stage larval (L1) lethal, while deletion heterozygotes developed normally, were fertile, and superficially similar to wild-type. Initial attempts to recover viable *atp-1*[R167Q] variant lines following CRISPR-Cas9 editing were unsuccessful as the variant animals were mostly embryonic lethal, even as heterozygotes. To facilitate the study of the R167Q and other proband variants, we first inserted an extra wild-type genomic *atp-1* copy into a safe harbor locus on chromosome IV (Fig. 3C) using the recombination-mediated cassette exchange method (RMCE) as previously described (Marom et al., 2023; Nonet, 2020). We reasoned that transgenic extra wild-type copies of *atp-1* would suppress the damaging effects of the variant alleles and allow them to grow as fertile adults.

RNA-seq analysis of homozygous *atp-1*[extra copy] animals showed that they express the transgenic copy at approximately half the level of the endogenous *atp-1* locus on chromosome I (Fig. S2A). Nevertheless, larval lethality of the homozygous endogenous *atp-1* locus deletion was fully rescued by the two copies of the wild-type transgenic *atp-1* on chromosome IV, allowing the animals to develop into fertile adults (Fig. S2B). This indicated that the transcriptional activity of the transgenic *atp-1* allele was sufficient to suppress the damaging effects of the variant alleles. Moreover, additional copies of *atp-1* in wild-type animals did not appear to be detrimental as homozygous *atp-1*[extra copy] animals were wild-type in terms of growth rate, body size, and crawling speed, although a minor decrease in thrashing speed was noted (Fig. S2C-F).

Using the *atp-1*[extra copy] lines, we were able to successfully generate viable animals harboring the proband variants R167Q, P316L, and S331F at the endogenous *atp-1* locus. In total, we generated two independent variant lines and one control line for each variant. The control lines are wild type at the variant residues, but harbor synonymous changes introduced during variant editing to generate a novel restriction site to aid in allele genotyping and prevent re-cleavage at the edited locus (Table S1A and S1B).

We first assessed the effects of *atp-1* variants on growth rate through post-embryonic development. Homozygous deletion (null), P316L, S331F, and R167Q variants without extra copies of *atp-1* died as embryos or arrested at the L1 stage. Therefore, we assessed growth rates of heterozygotes. At 72 hours post embryo lay, 100% of wild-type and control edits heterozygotes developed into healthy, fertile adults. However, most of the P316L and S331F heterozygotes only grew to L4 and L3 larval stages, respectively (Fig. 3D and E). In contrast, the majority (∼90%) of R167Q heterozygotes were embryonic lethal, and the remaining 10% grew to the L1 or L2 stages by 72 hours. No growth defects were observed in deletion (null) heterozygotes as 100% grew to mature adults by 72 hours, similar to wild-type and control edits (Fig. 3D and E). Together, these results indicated that the P316L, S331F, and R167Q variants are 1) damaging to *atp-1* function, 2) behave dominantly, and 3) are phenotypically different to the deletion (null) allele.

Although the P316L, S331F, and R167Q variant heterozygotes were markedly slow growing, some animals eventually developed into adults. This allows us to compare their body size and crawling speed using WormLab (Roussel et al., 2014). Body lengths of adults heterozygous for P316L, S331F, and R167Q were all significantly smaller (≤800µm) than wild-type, control edits and deletion heterozygotes (∼1000µm) (Fig. 3F). In addition, all three variant heterozygotes moved at a significantly slower crawling speed compared to their respective controls (Fig. 3G). Similar results were obtained for thrashing cycle (swimming) in liquid (Fig. S3A). Additionally, the absence of phenotypes in deletion heterozygotes indicates that *atp-1* is not a haploinsufficient gene in *C. elegans*. Based on these findings, we concluded that P316L, S331F, and R167Q are all change-of-function variants, possibly dominant negatives.

### Proband variants cause mitochondrial stress *in vivo*

Since *ATP5F1A* encodes a subunit of the mitochondrial ATP synthase (complex V), we hypothesized that proband variants altered mitochondrial function. To test this, we utilized a previously described mitochondrial unfolded protein response (UPR^mito^) reporter, *hsp-6p::gfp*, to measure mitochondrial stress (Taylor et al., 2014; Yoneda et al., 2004). Under normal conditions, the ATFS-1 transcription factor is imported into mitochondria and degraded.

However, under conditions of mitochondrial stress (e.g., low membrane potential, impaired protein import, accumulation of reactive oxygen species, unassembled subunits of the electron transport chain, or unfolded proteins), ATFS-1 does not enter the mitochondria and instead localizes to the nucleus where it facilitates the transcription of mitochondrial stress response factors, such as HSP-6, to help recover mitochondrial function (Fig. 4A) (Anderson & Haynes, 2020; Haynes et al., 2010; Rolland et al., 2019; Taylor et al., 2014; Yoneda et al., 2004). In wild-type, control edit and deletion heterozygotes, *hsp-6p::gfp* expression was barely detectable (Fig. 4B-G). However, in P316L, S331F, and R167Q heterozygotes, *hsp-6p::gfp* expression was strongly upregulated indicating significant mitochondrial stress, supporting the hypothesis that the variants disrupt mitochondrial function (Fig. 4B-G).

**Figure 4.**
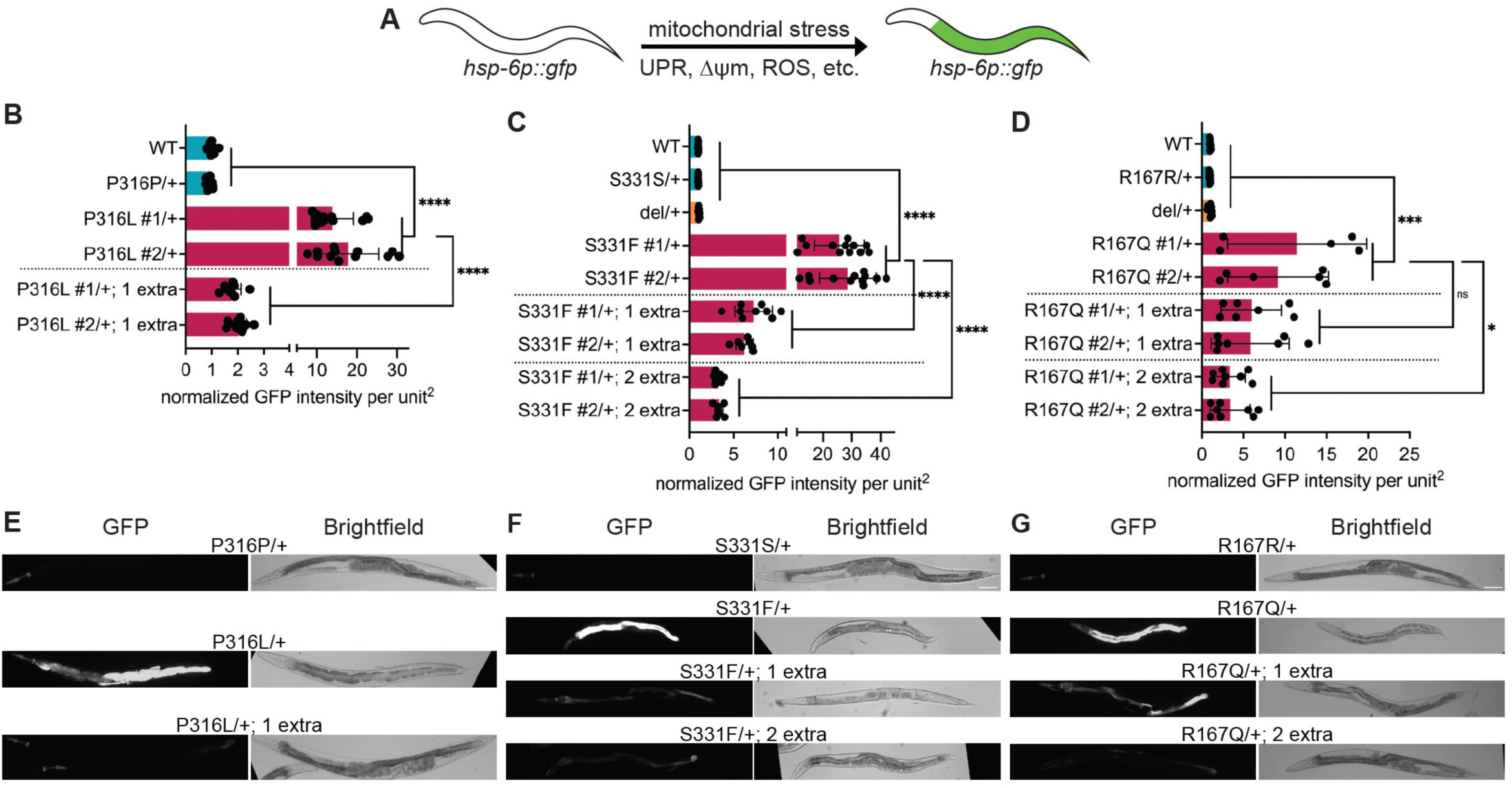
*atp-1* P316L, S331F, and R167Q variants induce mitochondrial stress. (A) Schematic of the *hsp-6p::gfp* reporter expressing strain. GFP is not expressed under normal conditions. However, upon mitochondrial stress, the *hsp-6p::gfp* reporter is transcriptionally upregulated in the intestine of the animal. (B-D) Normalized GFP fluorescence expression in 1- day adult animals. (E-F) Representative images of animals quantified in B-D. Scale bar, 100µm. See Fig S7 for crossing scheme to generate these animals. ns – not significant, * p<0.05, ** p<0.01, *** p<0.001, **** p<0.0001.

### Proband variants act through a dominant negative mechanism

Dominant phenotypes observed with the *atp-1* variants, but not the deletion (null) (Figs. 2 and 3), as well as the dominant presentation in the *ATP5F1A* probands suggest a change-of-function genetic mechanism. From classic gene dosage studies, if the genetic mechanism is antimorphic/dominant negative, then the addition of wild-type transgenic copies should suppress the phenotype (Muller, 1932). The mitochondrial stress phenotype (i.e., induction of *hsp-6p::gfp* expression) was completely suppressed by the addition of one extra copy of wild-type *atp-1* for P316L (Fig. 4B and E). Addition of one extra wild-type copy of *atp-1* partially suppressed mitochondrial stress for S331F and R167Q, and addition of two extra copies further suppressed mitochondrial stress to close to wild-type levels (Fig. 4C, D, F and G). The absence of a phenotype in deletion (null) heterozygotes and the suppression of mitochondrial stress in variant heterozygotes by the addition of extra copies of the wild-type allele provides evidence that the P316L, S331F, and R167Q variants act as dominant negative alleles.

### Proband variant alters mitochondrial morphology *in vivo*

The dynamics of mitochondrial fission/fusion can change in animals undergoing mitochondrial stress to help maintain ATP homeostasis (reviewed in (Liu et al., 2020) and (Campbell & Zuryn, 2024)). For example, during *C. elegans* starvation, mitochondria are known to fuse to help produce more ATP (Gomes et al., 2011; Liesa & Shirihai, 2013; Rambold et al., 2011; Tondera et al., 2009; Weir et al., 2017). Conversely, under nutrient-rich conditions, mitochondria undergo fission in an effort to decrease ATP production (Molina et al., 2009). To evaluate the effect of the proband variant on mitochondrial morphology, a transgene expressing a mitochondrial-targeted GFP (mito-GFP) was used to label the mitochondria in the body wall muscles (Fig. 5A) (Rolland et al., 2013). High-resolution images were acquired by confocal microscopy, and image analysis was performed using Mitochondrial Segmentation Network (MitoSegNet), a pretrained deep learning segmentation model for quantifying mitochondrial morphology (Fischer et al., 2020). Since all three variants induced mitochondrial stress, we performed these studies only in the P316L variant. In wild-type and P316P control muscle cells, mitochondria were elongated tubules and were organized largely as parallel arrays that extend along the thin filaments (Fig. 5A) (Schultz et al., 2017). In the P316L variant muscle cells, the average mitochondrial area was increased (Fig. 5A and B). In addition, the width of the minor tubule axis was also increased (Fig. 5B), giving a more rounded or globular morphology (Fig. 5A and B). No difference in the long (major tubule) axis was noted between P316L variant and P316P control (Fig. S3B), suggesting that mitochondrial fragmentation did not account for this difference. The mitochondria of the deletion heterozygotes were not different from the control heterozygotes (Fig. 5A and B). These results indicate that the mitochondrial morphology is altered in the P316L variant, and likely also in the R167Q and S331F variants.

**Figure 5.**
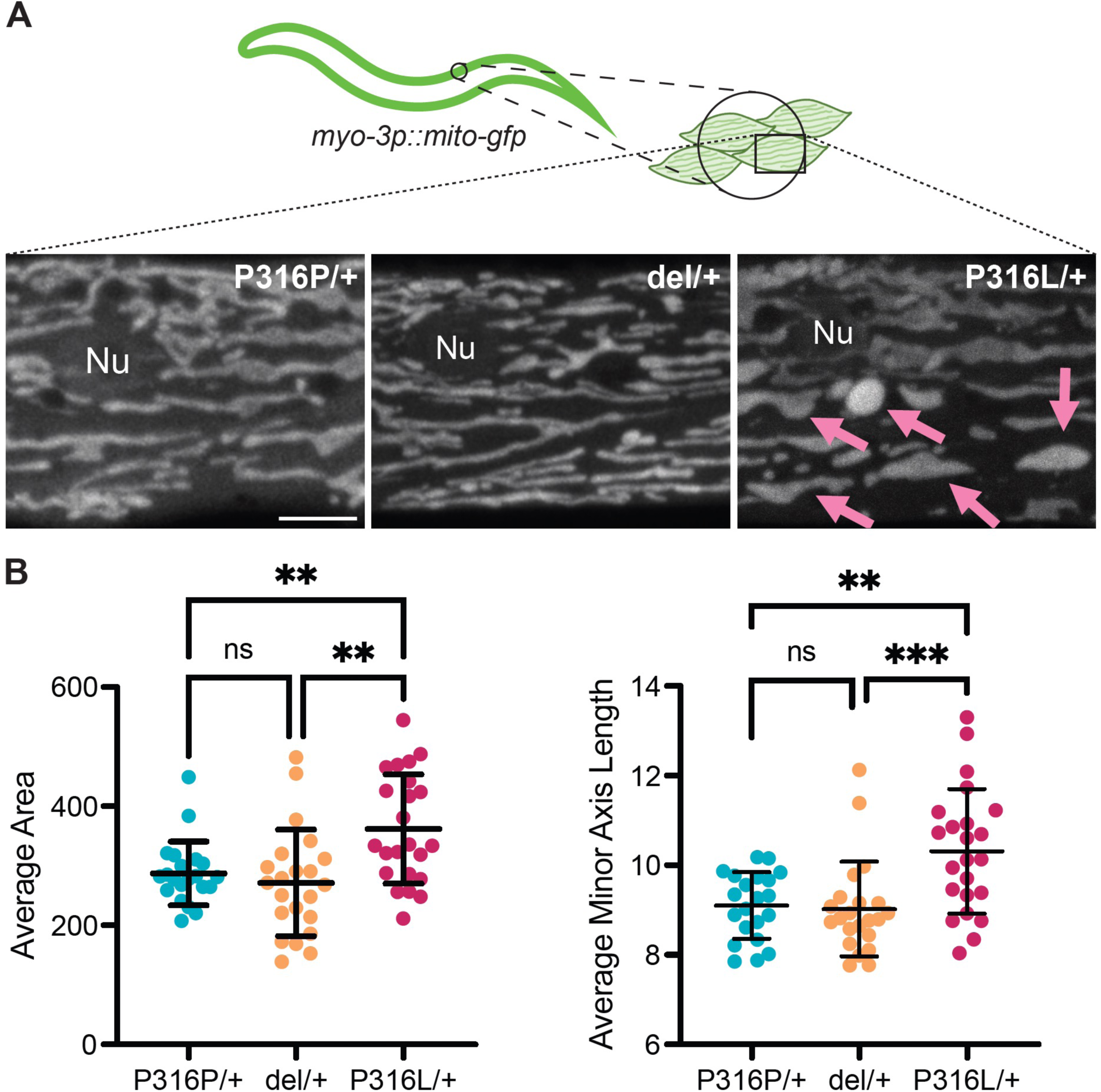
*atp-1* P316L variant alters mitochondrial morphology. Mitochondrial morphology is altered in P316L animals. (A) Representative images of mitochondrial marker *bcIs80* (*myo- 3p::mito-gfp*) expressed in the body wall muscles. Scale bar, 5µm. (B) Quantification of average area and minor axis length per animal, calculated using MitoSegNet. 21-23 animals imaged per genotype. ns – not significant, ** p<0.01, *** p<0.001, **** p < 0.0001.

### Proband variants, p.Arg182Gln, p.Ser346Phe and p.Leu109Ser, disrupt ATP synthase (Complex V) enzyme activity

Next, we evaluated mitochondrial function in skin fibroblasts from probands 1, 4, and 6 representing the variants p.Arg182Gln, p.Ser346Phe, and p.Leu109Ser, respectively (Fig. 6A). Fibroblasts from the other probands were not available for study. On blue native PAGE, complex V migrated as a single band in all three probands as in the controls. Compared to 50 control fibroblast lines, the in-gel activity of complex V was markedly reduced in proband 6 (p.Leu109Ser), moderately reduced in proband 1 (p.Arg182Gln) and very mildly reduced in proband 4 (p.Ser346Phe) (Fig. 6B). In-gel activities of other complexes (I, II, and IV) were normal. This pattern in *ATP5F1A* fibroblasts is different than that noted for *ATP5PO* or in *MT*- *ATP6* variants where in the setting of Coomassie blue stained gels, the separation of the peripheral stalk results in complex V bands of lower MW (Ganapathi et al., 2022; Smet et al., 2009; Wittig et al., 2010), whereas for *ATP5F1A* probands, similar to *ATP5F1D* probands, the catalytic activity is reduced at the intact complex V without lower MW bands (Ganapathi et al., 2022; Olahova et al., 2018).

**Figure 6.**
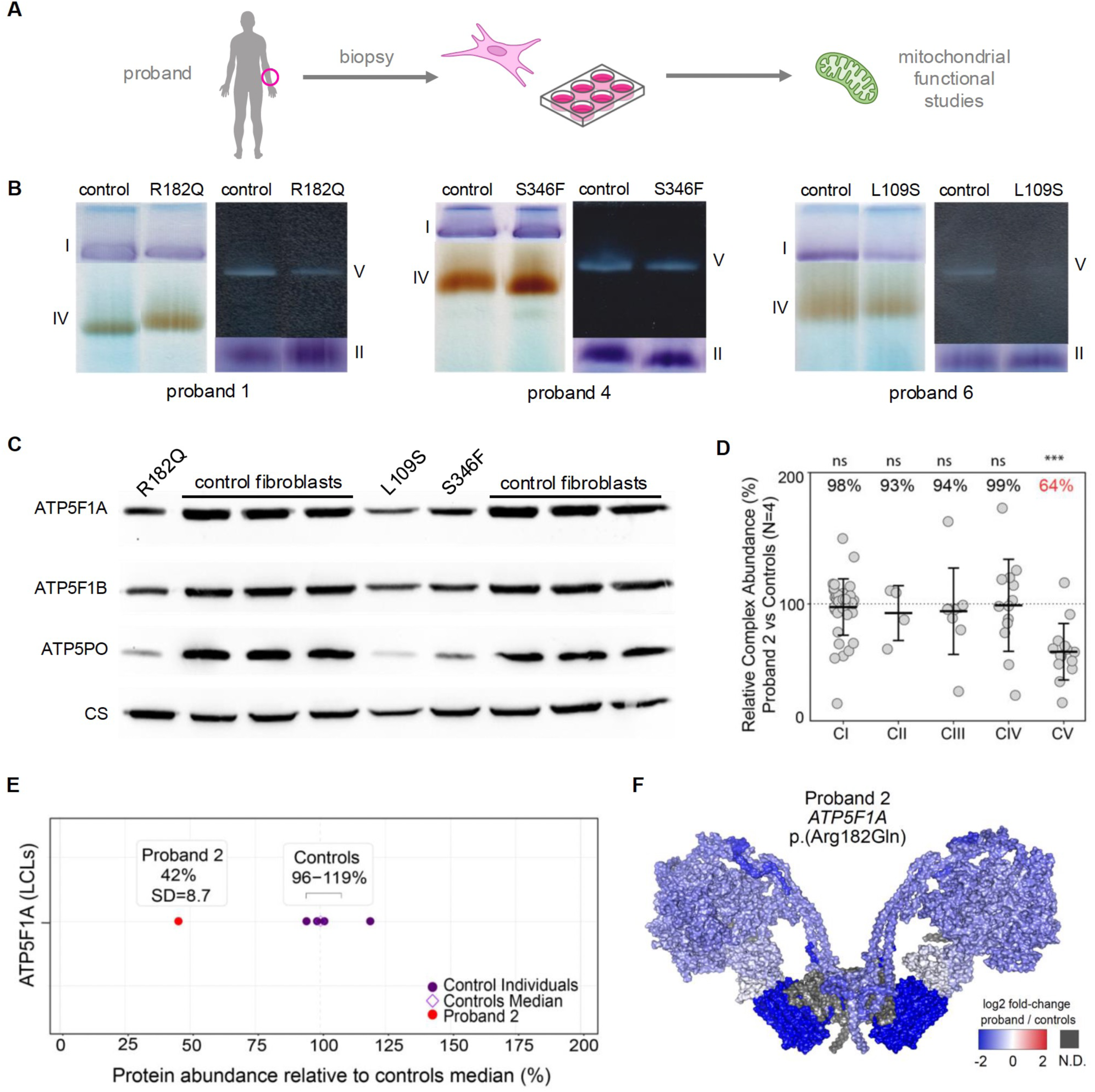
**Proband fibroblasts show reduced complex V abundance and activity**. (A) Schematic of mitochondrial functional studies performed on proband fibroblasts. (B) BN-PAGE analysis of complexes of the electron transport chain of proband 1 (p.Arg182Gln), proband 4 (p.Ser346Phe) and proband 6 (p.Leu109Ser) fibroblasts. (C) Western blots analysis of ATP5F1A, ATP5F1B and ATP5PO of proband and control fibroblasts. cs, citrate synthase, loading control. (D) Relative Complex Abundance (RCA) of OXPHOS complexes from proteomics data of Proband 2 (p.Arg182Gln) LCLs compared to controls (N=4) showing an isolated Complex V defect. Middle bar represents mean complex abundance. Upper and lower bars represent 95% confidence interval. Significance was calculated from a two-sided t-test between the individual protein means. *** = p<0.001, ns = not significant, p>0.05. (E) Protein range for ATP5F1A in LCLs of Proband 2 (p.Arg182Gln) (red dot) and controls (N=4, purple dots) showing a standard deviation of 8.7 from the control median. (F) Topographical heatmap of the log2 fold-change abundances of Proband 2 relative to controls onto the cryo-EM structure of the dimer complex V structure (PDB: 7AJD).

The complex V ATPase hydrolytic enzyme activity in fibroblasts was decreased in all three variant cell lines at 42% (p.Arg182Gln), 33% (p.Ser346Phe), and 37% (p.Leu109Ser) of the average of control cells (Table S2). These enzyme activities were similar to those previously reported for the dominant p.Arg207His variant (Lines et al., 2021; Zech et al., 2022), but more than those reported for probands with the recessive p.Arg329Cys variant (29% and 19%, R. Rodenburg, personal communication) (Jonckheere et al., 2013).

Western blot analysis of different complex V subunits indicated that ATP5F1A (α- subunit) and ATP5F1B (the β-subunit that associates with ATP5F1A) were moderately reduced while ATP5PO (the OSCP-subunit that also associates with ATP5F1A and ATP5F1B), was strongly reduced in all three probands (Fig. 6C).

Mass spectrometry based proteomic analysis of proband 2 (p.Arg182Gln) lymphoblastoid cells (LCLs) revealed a reduction of almost all complex V subunits (64% control mean) as illustrated in relative complex abundance (RCA) analysis (Fig. 6D and S4A). Consistent with the western blot data, the abundance of the ATP5F1A protein in proband 2 was reduced to 42% of in assay controls median (Fig. 6E). On blue native PAGE, complex V migrated as a single markedly reduced band in proband 2’s LCLs (Fig. S4B), similar to proband 1 fibroblasts carrying the same p.Arg182Gln variant (Fig. 6B). Likewise, proteomic analysis of fibroblasts of probands 1, 4 and 6 revealed a comparable decrease in the RCA of complex V in all three probands (53-58%), and in the RCA of complex V over complex II (Table 2). There was a mild increase in the RCA of complex III. Similar to western blot findings, the ATP5F1A protein was reduced to 44-48% of controls whereas the ATP5PO protein in proband 1, 4, and 6 was reduced to 34-39% of controls (Table 2).

**Table 2.** Respiratory chain complex abundances in fibroblasts of patients with complex V deficiency and controls.

| Gene | Proband | Relative complex abundance |  |  |  |  | RCA ratios |  |  | Relative protein levels |  |  |
| --- | --- | --- | --- | --- | --- | --- | --- | --- | --- | --- | --- | --- |
|  |  | CI | CII | CIII | CIV | CV | CI/CII | CIV/CII | CV/CII | ATP5F1A | ATP5F1D | ATP5PO |
| *ATP5F1A* | 1 | 112 | 106 | **128** | 95 | **58** | 1.057 | 0.896 | **0.547** | **48%;-7.5** | 58%;-2.5 | 39%;-7.2 |
| *ATP5F1A* | 4 | 110 | 97 | **132** | 115 | **53** | 1.134 | 1.186 | **0.546** | **44%;-8.4** | 51%;-3.1 | 34%;-8.6 |
| *ATP5F1A* | 6 | **115** | 102 | **138** | 117 | **58** | 1.127 | 1.147 | **0.569** | **44%;-8.4** | 69%;-1.6 | 34%-8.4 |
| *ATP5F1D* |  | 101 | 101 | 110 | 105 | **32** | 1.000 | 1.040 | **0.317** | 19%;-18.0 | **32%;-5.7** | 16%;-15.3 |
| *ATP5PO* |  | 101 | 104 | **124** | 103 | **56** | 0.971 | 0.990 | **0.538** | 95%;+0.3 | 103%;+0.6 | **13%;-16.9** |
| **Controls N=23** |  |  |  |  |  |  |  |  |  |  |  |  |
| Mean±SD |  | 97.4±7.3 | 97.1±9.4 | 100.3±6.3 | 99.6±15.1 | 96.5±7.5 | 1.01±0.12 | 1.03±0.20 | 1.03±0.20 |  |  |  |
| Range |  | 87-114 | 77-110 | 88-114 | 83-150 | 84-114 | 0.84-1.36 | 0.83-1.79 | 0.81-1.36 | 76-110% | 65-97% | 69-116% |

The relative complex abundances of the respiratory chain complexes and their internal ratios, and the relative amounts of the individual proteins as determined by proteomics analysis of fibroblasts after correction for mitochondrial abundance are listed as percent of in-assay controls (n = 5) for our probands. These are shown in comparison to a range of 23 normal controls and to cell lines of patients with other nuclear encoded autosomal recessive complex V defects. The mean, the standard deviation (SD) and the observed range among controls are provided. The relative protein levels are shown as percent of in assay controls; and as the Z-score of the distribution observed in normal control population. This control data did not deviate significantly from a normal distribution. Patient results significantly outside the range observed in control subjects are shown in bold. Abbreviations: CI = complex I, CII = complex II, CIII = complex III, CIV = complex IV, CV = complex V. C/CII is the ratio of the RCA of complex I divided by the RCA of complex II. The RCAs of the large subunit of the mitoribosome, the small subunit of the mitoribosome and the pyruvate dehydrogenase complex were all normal and are therefore not shown.

Since *ATP5F1A* is expected to be tolerant to loss-of-function variants (pLI=0.29; o/e=0.3 – 0.58), a ∼50% reduction in abundance of ATP5F1A is not predicted to be damaging. Therefore, the marked reduction in complex V enzyme activity (below 50% that of controls) and the observed reduction in abundance of both ATP5F1A-interacting F1 and distal Fo complex V subunits (as reflected in Fig. 6F and S4C) indicates a destabilizing effect of the proband variants on complex V assembly and function, consistent with a dominant negative mechanism.

Interestingly, dominant negative *ATP5F1A* proband fibroblasts or LCLs showed a milder reduction in complex V subunits when compared to fibroblasts of probands with biallelic loss of function variants in *ATP5F1D* (Olahova et al., 2018) and *ATP5PO* (Ganapathi et al., 2022).

### Decreased ATP and mitochondrial membrane potential despite increased oxygen consumption suggests uncoupled oxidative phosphorylation

To better understand the pathogenic mechanism of the *ATP5F1A* variants, we measured oxygen consumption, mitochondrial membrane potential, and steady state ATP in cells. On the Seahorse 96XF bioanalyzer (Agilent) mitochondrial stress test comparison of the proband 1 p.Arg182Gln variant fibroblasts and age-, sex-, and race-matched control fibroblasts, the proband 1 fibroblasts displayed significantly higher rates of basal respiration (i.e., unstressed) and an overall higher capacity for maximal respiratory function (Fig. 7A and B, and Fig. S5). As a result, the spare respiratory capacity in proband fibroblasts was also overall higher, albeit not significant, compared to wild-type fibroblasts (Fig. 7B and Fig. S5). Despite the increased rates of oxygen consumption, the membrane potential of proband fibroblast mitochondria measured by the fluorogenic dye, tetramethylrhodamine ethyl ester (TMRE), was significantly lower than that of wild-type control (Fig. 7C) and was as low as that of wild-type fibroblasts treated with the uncoupling agent carbonyl cyanide-p-trifluoromethoxyphenylhydrazone (FCCP) (Fig. 7C).

**Figure 7.**
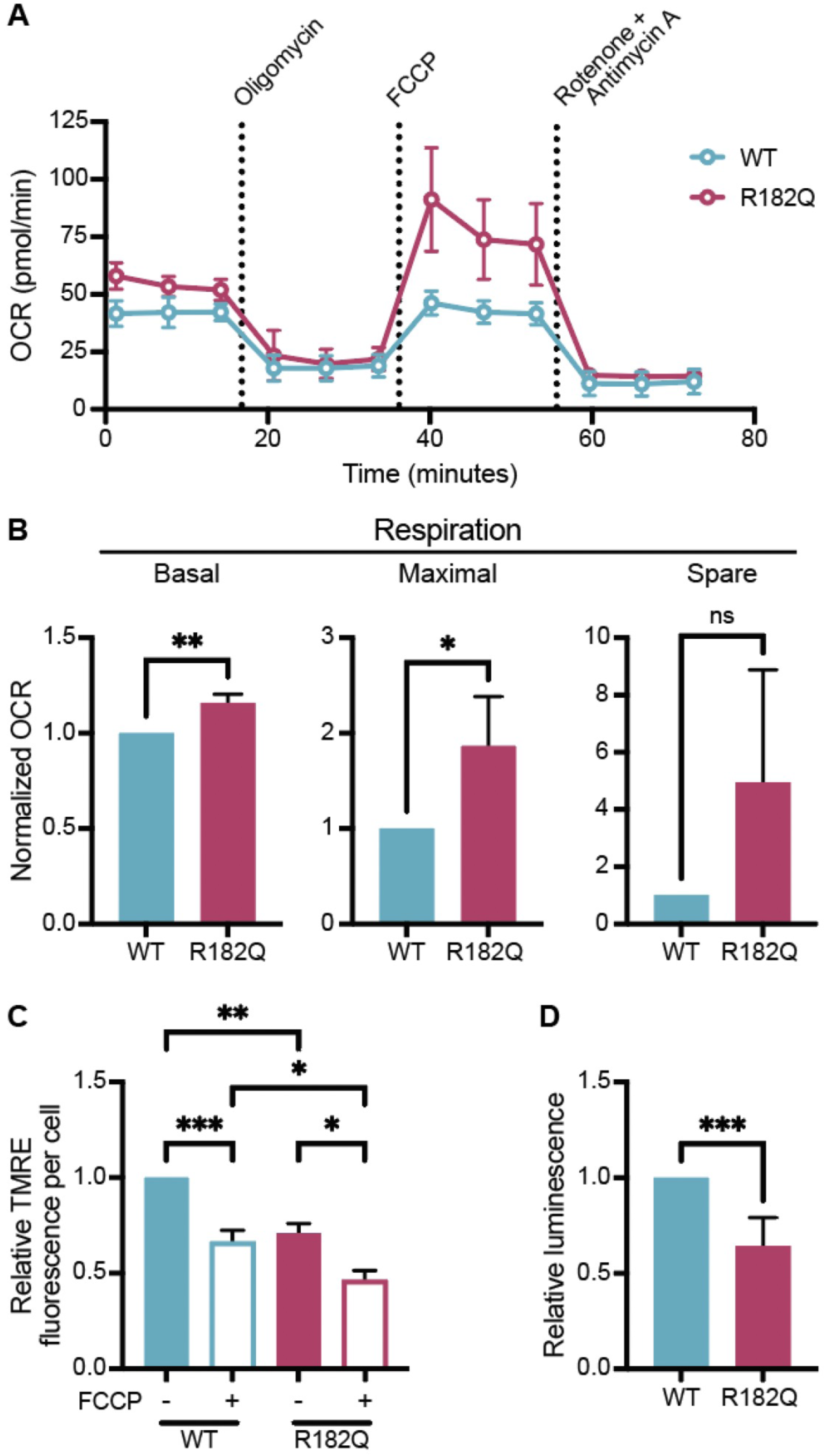
*ATP5F1A* p.R182Q proband fibroblasts show reduced ATPase activity and uncoupled oxidative phosphorylation. (A) Representative oxygen consumption rate (OCR) of proband derived fibroblasts compared to age, race, and sex matched control fibroblasts measured by Seahorse in intact cells. (B) Normalized average basal, maximal, and spare OCR from three biological replicates. Individual graphs from each experiment are shown in Fig. S6. (C) Membrane potential of proband and control fibroblasts as measured by TMRE fluorescence with and without FCCP treatment. (D) Total cellular ATP content taken from lysed proband and control fibroblasts using a luciferase reporter. ns – not significant, * p <0.05, ** p<0.01, *** p<0.001, **** p < 0.0001.

Furthermore, the proband fibroblasts exhibited significantly diminished ATP steady state levels compared to wild type controls (Fig. 7D). These results are opposite to those reported for the p.Arg207His variant where the cells exhibited a lower rate of oxygen consumption and no change in mitochondrial membrane potential compared to control cells (Lines et al., 2021).

Collectively, these results indicate that in fibroblasts the inefficient ability of the *ATP5F1A* p.Arg182Gln variant to generate ATP is mostly the result of uncoupled oxidative phosphorylation. Cells use the normal activities of complexes I through IV resulting in higher oxygen consumption, but their defective complex V results in loss of membrane potential and decreased ATP.

## DISCUSSION

We describe a series of 6 new probands with 4 heterozygous *de novo* missense variants in *ATP5F1A*, who presented with intellectual disability, developmental delay, prominent motor symptoms with variable degrees of dystonia and spasticity. Structural analysis suggested that the four variants likely disrupt the ATP5F1A’s (α-subunit) interactions with the β− (ATP5F1B) and/or γ− (ATP5F1C) subunits of ATP synthase. Functional studies in *C. elegans* determined that all variants evaluated were damaging and act by a dominant negative mechanism. Analysis of proband-derived cells showed reduced ATPase activity and decreased abundance of ATP5F1A and other complex V subunits. For two proband variants, uncoupled oxidative phosphorylation was likely a mechanism of disease. Our study brings the total described families with dominant *de novo* missense variants in *ATP5F1A* currently to 12, making this one of the most common causes of nuclear genome encoded complex V deficiency together with recessive *TMEM70* variants (Honzik et al., 2010; Magner et al., 2015). Furthermore, except for *ATP5F1A* and *ATP5F1B*, autosomal recessive mode of inheritance is currently the only known genetics for complex V deficiency associated with *ATPAF2, ATP5F1E, ATP5PO, ATP5MD, ATP5F1D* and *TMEM70* (OMIM). Thus, the autosomal dominant mode of *ATP5F1A* pathogenicity, and likely also for *ATP5F1B*, may have been previously underappreciated. From a clinical diagnostic perspective, this study also illustrates that if a variant of uncertain significance is identified in *ATP5F1A*, multiple methods can potentially confirm the pathogenicity including: 1) blue native PAGE with in-gel activity staining, 2) hydrolytic ATPase complex V enzyme assay, 3) proteomics studies to identify both the reduced abundance of complex V via RCA and of the ATP5F1A protein, 4) mitochondrial respiration (Seahorse) analysis, and, 4) functional studies in model organisms. It is worth noting that the reduction in abundance of ATP5F1A and other complex V proteins observed in this study is similar to what is observed in variants in other parts of the catalytic central globular head of complex V, such as in *ATP5F1D*, and that identification of the correct molecular cause remains paramount for a final diagnosis (Fig. S4C). On the other hand, the impact of dominant negative *ATP5F1A* variants on complex V subunit abundance is clearly different from that of recessive variants in *ATP5F1D* and *ATP5PO* (Fig. S4C), where the central stalk remains preserved.

Although so far rare in mitochondrial complexes, a dominant negative effect can be observed in proteins that form hetero-multimers and can involve disrupting interactions between subunits (Anders & Botstein, 2001; Bejsovec & Anderson, 1988; Kemphues et al., 1980; Lund et al., 1997). The catalytic head domain of ATP synthase consists of 3 α- and 3 β-subunits, which interact with the central stalk (γ-subunit). Protein modeling of ATP synthase showed that all four residues, Arg182, Ser346, Pro331, and Leu109, affected in the probands are located at or near the contact points of α:β and/or α:γ subunits of the F_1_. Therefore, substitution of these residues with the proband variant is predicted to alter the protein conformation and disrupt the interactions between the α−, β− and/or γ−subunits leading to altered ATP synthase function.

Indeed, our BN-PAGE, western blot, electron-transport chain enzyme assays, and proteomic analysis showed reduced activity and abundance of complex V subunits. Since *ATP5F1A* is not a haploinsufficient gene (pLI=0.29, gnomAD v4.1), the observed complex V defects cannot be explained simply by ∼50% reduction in function or abundance of ATP5F1A. We postulate a dominant negative mechanism whereby the proband variants negatively affect complex V and alter its function and stability. Our hypothesis is based on the supposition that 1/8^th^, 3/8^th^, 3/8^th^ and 1/8^th^ of the complex V consists of 3, 2, 1 and 0 variant α-subunits, respectively (see Fig. S6). We propose that complexes having 2 or 3 variant α-subunits (4/8^th^ or 50%) are likely unstable and undergo degradation while 3/8^th^ of the complex comprising of only 1 variant α- subunit is stable but has reduced function. The ATP5PO, which interacts with both the α-subunit and the β−subunit, was strongly reduced with impact on the stability of the side chain. The remaining 1/8^th^ of the complex comprising of all wild-type α-subunits functions normally. Thus, the variable levels of ATP5F1A and complex V observed in our probands likely reflects the variable activity, stability and dominant negative effect of the variant protein on the conformation of the assembled complex V. A similar dominant negative effect was recently postulated for a variant in *ATP5F1B*, which encodes the β-subunit of ATP synthase (Ganetzky et al., 2022; Nasca et al., 2023). However, in contrast to *ATP5F1B* probands (Ganetzky et al., 2022), the *ATP5F1A* probands described here showed no clinical evidence of hypermetabolism. Of note, the dominant negative *de novo ATP5F1A* cases have less severe clinical presentation than the homozygous cases (Jonckheere et al., 2013; Lieber et al., 2013). This is likely because in the dominant negatives cases, 1/8^th^ of complex V which comprises of all wild-type α-subunits is fully functional.

Dominant *de novo* missense variants in *ATP5F1A* have previously been reported, however, functional evaluation has only been described for one variant, p.Arg207His (Lines et al., 2021; Zech et al., 2022). Four individuals with this variant presented with life-threatening, neonatal onset mitochondriopathy with failure to thrive, hyperlactatemia and hyperammonemia. Interestingly, all four showed spontaneous improvement and clinical resolution in late infancy with no persistent neurological phenotypes. Biochemical analysis of p.Arg207His fibroblasts showed reduced complex V ATPase activity (<20%-30% of controls), reduced ATP5F1A subunit protein abundance (Lines et al., 2021; Zech et al., 2022), but lower oxygen consumption rate with no change in mitochondrial membrane potential (Lines et al., 2021). All probands in our study also showed a similar reduction in ATPase activity (∼33-42% of controls) and reduced abundance of ATP5F1A and other complex V subunits. However, in contrast to p.Arg207His proband, our p.Arg182Gln proband (proband 1) fibroblasts showed higher basal and maximal respiration rates and reduced mitochondrial membrane potential suggesting uncoupled oxidative phosphorylation. Although the exact molecular mechanism remains to be determined, the uncoupled oxidative phosphorylation is similar to that reported for the dominant negative variant in *ATP5F1B* (Ganetzky et al., 2022), thus representing a shared mechanism for dysfunction in the catalytic head, that is distinct from that of *MT-ATP* variants. Moreover, the distinct clinical presentations and mitochondrial physiology findings between p.Arg207His individuals and our p.Arg182Gln proband suggests that dominant variants in *ATP5F1A* can cause disease via additional distinct genetic and molecular mechanisms.

Additional support for uncoupled oxidative phosphorylation in our probands comes from our *C. elegans* studies that showed altered mitochondrial morphology. The elongated tubular mitochondria of the wild-type body wall muscle cells became more rounded and globular in the *atp-1*[P316L] variant animals. Interestingly, cultured cells treated with compounds that uncouple the mitochondria or cause loss of mitochondrial membrane potential displayed similar changes in mitochondrial morphology from a tubular to a globular shape (Miyazono et al., 2018). Although the changes observed in cultured cells may not be directly comparable to those in *C. elegans* muscle cells, these results lend further support for mitochondrial uncoupling in *atp- 1*[P316L] mitochondria. Of note, *atp-1*[P316L] corresponds to *ATP5F1A* p.Pro331Leu (proband 5) and *atp1* P328L in yeast, a known mitochondrial genome integrity (*mgi*) mutant (Chen & Clark-Walker, 1995, 1996; Clark-Walker et al., 2000; Wang et al., 2007). All *mgi* mutants identified so far are without exception, dominant, map to α, β, or γ subunits of ATP synthase, and uncouple oxidative phosphorylation (Wang et al., 2007). These observations suggest that oxidative phosphorylation is also uncoupled in our p.Pro331Leu (proband 5) variant and indicate that uncoupled oxidative phosphorylation may be a common feature in our probands. Analogous findings were recently reported by Ganetzky et al., who described a dominant negative, uncoupling, missense *ATP5F1B* variant in monozygotic twins with a higher oxygen consumption rate and lower mitochondrial membrane potential (Ganetzky et al., 2022). Interestingly, the *ATP5F1B* variant is located near a known *mgi* locus, and the twins have failure to thrive and developmental delay, similar to the probands in this study. Therefore, yeast conserved legacy *mgi* mutants could provide helpful functional data in determining the pathogenicity and genetic mechanism of ATP synthase variants that may in the future be identified in human patients.

In summary, using an integrated multidisciplinary approach which included structural modeling, genetic and cell biology studies in *C. elegans*, and biochemical and mitochondrial physiology studies in patient cells, we provide strong evidence that *de novo* heterozygous variants, p.Arg182Gln, p.Ser346Phe, p.Pro331Leu, and p.Leu109Ser, in *ATP5F1A* are damaging and function via a dominant negative mechanism. Comparison of our cases with others reported in the literature suggested that not all dominant *de novo ATP5F1A* variants work via the same genetic and/or molecular mechanism, underscoring the importance of *in vivo* and *in vitro* functional analyses for a correct molecular diagnosis. There may also be differences in pathophysiological mechanism between cell types, as pertinent differences between fibroblasts and neuronal cells have been reported in some complex V deficient patients (Lorenz et al., 2017). We recommend that a similar integrated approach be used for characterizing future novel variants in *ATP5F1A* and other ATP synthase subunits to obtain a more complete understanding of the variant mechanism, which is a prerequisite for developing an effective treatment strategy. There are now 12 individuals reported with *de novo* heterozygous variants in *ATP5F1A,* making *ATP5F1A* the most frequent nuclear genome cause of complex V (ATP synthase) deficiency. Our study expands the spectrum of *ATP5F1A*-associated conditions to include dominant negative variants that likely uncouple oxidative phosphorylation and ATP production.

## MATERIALS & METHODS

### Consent for genetic study

Families of probands 1 and 5 provided informed consent for participation in the Undiagnosed Diseases Network (UDN), under a protocol approved by the NIH IRB (15HG0130). Proband 2 was enrolled in the Rare Disease Now Program at the Murdoch Children’s Research Institute (HREC 67401) (Australia) and parents provided informed consent for genome sequencing, proteomic studies and publication of clinical and molecular data. Written informed consent was obtained from participants and/or their legal guardians for proband 3 for the collection, research use, and storage of the specimens according to the protocol approved on July 1, 2020, by Indiana University. Fibroblast studies in Colorado were studied under a protocol approved by the Colorado Multiple Institutional Review Board COMIRB protocol # 18-1828, and probands 1 and 4 were further studied in Colorado under COMIRB protocol # 16-0146 after obtaining informed consent. Clinical de-identified data of Proband 6 was analyzed with IRB approval under Baylor College of Medicine study protocol (H-48670), and the fibroblast studies were ordered as part of his clinical care.

### Sequencing and bioinformatic analysis

Trio exome or whole genome sequencing was performed for probands 1, 5 and 6 by Baylor Genetics as described previously (Keehan et al., 2021). Variant filtering was performed using Codified Genomics software. Variants with an allele frequency of > 0.01 in gnomAD (v.2.1.1) were filtered out. *De novo*, compound heterozygous, and X-linked single nucleotide and small indel variants were prioritized for review. Trio WGS for proband 2 was processed by the Centre for Population Genomics (Australia) using the DRAGEN GATK best practices pipeline. Reads were aligned to the hg38 reference genome with Dragmap (v1.3.0), and joint variant calling across the cohort was performed using GATK HaplotypeCaller (v4.2.6.1) for SNVs and indels. Variants were annotated with VEP 110 and integrated into Seqr (Pais et al., 2022) for filtration and analysis. For patients 3 and 4, initial clinical exome sequencing was performed by GeneDX (Gaithersburg, MD), identifying a heterozygous variant of uncertain significance (VUS) in ATP5F1A [(NM_004046.6:c.1037C>T, p.(Ser346Phe)]. For patient 3, subsequent parental testing confirmed that this variant arose *de novo*. To further investigate the genetic findings, research-based whole genome sequencing (WGS) data re-analysis was conducted as described (Jacobs et al., 2022) using the EMEDGENE analytical pipeline (https://iuhealth.emedgene.com/) and supplemented with manual curation. Identified variants were filtered based on minor allele frequency (<0.1% in gnomAD), conservation, and deleteriousness by *in silico* prediction tools (PolyPhen-2, MutationTaster, REVEL, DANN, LRT, and SIFT).

### 3D Molecular Modeling

To explore the impact of the missense variants identified in this study we employed 3D molecular modeling as described (Booth et al., 2020; Booth et al., 2024). Briefly, previously described structural models comprising the 10-subunit human ATP synthase (PDB: 8H9S), were obtained (Lai et al., 2023). Alignment, visualization, and mutagenesis were performed by using PyMOL (version 2.5.5). Note single letter amino acid abbreviations were used to denote amino acids of interest.

### C. elegans strains and culturing conditions

Single letter amino acid abbreviations were used to denote *atp-1* variants studied in *C. elegans.* P316L and S331F variant lines and corresponding controls (P316P and S331S) were maintained and all assays for these lines were performed at 20° C on NGM plates, while R167Q variant and R167R control lines were maintained and all assays except the development assay (Fig. 3D) were performed at 15° C as the strong embryonic and larval lethality of this variant was partially suppressed at this temperature. Strains used in this study were as follows: *tmC5* (balancer for the *atp-1* extra copy on IV), *hIn1* (balancer for *atp-1* deletion and variants), *bcIs80* [*myo-3p::mito-gfp* + pRF4] (a generous gift from Barbara Conradt, unpublished), *zcIs13* [*hsp- 6p::gfp*], and *atp-1(ok2203)*. A full list of strains their genotypes are listed in Table S3.

### Transgene generation

Using Recombination Mediated Cassette Exchange (RMCE), an extra wild-type copy of *atp-1* was inserted as a single copy onto chromosome IV (Nonet, 2020). This extra copy had a modified PAM sequence used for the R167 variant and control edits so that Cas9 would only edit the endogenous copy of *atp-1 and* had an additional silent NciI restriction site added in order to differentiate the extra copy on IV from the endogenous copy on I in RNA-seq (Table S4). To generate the P316 and S331 variants, the extra copy was first edited using the corresponding control edit repair template to destroy the PAM for each locus, and subsequently the newly generated variant edits and control edits at the endogenous locus were crossed back to the original extra copy line.

Since all variant edited strains were homozygous lethal, they were maintained over a balancer chromosome (see Fig. S7). Due to the infertility of the heterozygotes, all variant strains were also maintained with one extra copy of *atp-1*, over a second balancer chromosome. The extra copy and balancer for the endogenous copy were removed by crossing prior to phenotyping (Fig. S7).

### RNAseq and Analysis

To determine the relative expression level from the *atp-1* extra copy transgene compared to the endogenous *atp-1* gene, RNA-seq was performed with strain udnSi40 (Table S3), which is homozygous for both *atp-1* loci, followed by read analysis as previously described (Marom et al., 2023). Two single nucleotides silent polymorphisms in the *atp-1* extra copy transgene were used to distinguish transcripts from the two *atp-1* loci. While the *atp-1* transgenic extra copy was found to be expressed at about 50% the level of the endogenous *atp- 1* gene (Fig S2A), the wild-type extra copy gene can still be used in gene dosage studies to assess whether one or two additional wild-type copies can suppress the dominant, change of function, *atp-1* variant alleles.

### CRISPR-Cas9 gene editing

CRISPR-Cas9 editing was performed as previously described (Fielder et al., 2022; Huang et al., 2022). Before knocking in the P316L and S331F variants, we first introduced two silent base pair changes to the transgenic *atp-1* extra copy to facilitate CRISPR-Cas9 editing of the endogenous *atp-1* locus and to prevent editing of the transgenic copy. Briefly, edited *atp- 1*[extra copy] animals 24 hours post larval L4 stage were injected with Cas9 protein (IDT 1081060), tracrRNA (IDT 1072533), pRF4 plasmid as a positive injection control, and IDT Alt-R® CRISPR-Cas9 sgRNA (Table S1A) and repair template (Table S1B). Repair templates were 100 bp in length, centered around the proband variant. Silent changes were made to eliminate the PAM, and to add or eliminate a restriction site (FokI eliminated in R167, XbaI added in S331, and HaeIII added in P316). To ensure any phenotypes observed were due to the proband variants and not the silent changes, control repair templates were used that only contained silent changes (referred to in text as P316P, S331S, and R167R). Variant edit lines were crossed back to the original extra copy line so that all lines were maintained with the same extra copy.

### Development assays

L4 double balanced heterozygotes with one extra copy were mated with wild-type males for 24 hours at 20° C (Fig. S7). Next, mated animals were allowed to lay embryos on an empty plate for 2 hours and then removed. 40 Embryos for controls or 90 embryos from variants were moved to a separate plate and allowed to grow at 20° C for 72 hours. 24 hours after embryo laying, remaining embryos on plates were scored as dead embryos. At 72 hours post embryo laying, stages of animals and genotypes were quantified. Four replicates were performed. For R167Q, 35-70 embryos were used as mated animals laid less than 90 total embryos.

### Length, crawling, and thrashing assays

Wormlab experiments were performed as previously described (Huang et al., 2022). Briefly, double heterozygous animals were mated to VC2010 males and progeny allowed to grow at 20° C or 15° C (R167 group) (Fig. S7). All animals assayed were heterozygous for *tmC5* balancer, aside from VC2010 control (Fig. S7). 1-day-old adult progeny were gently transferred to an NGM plate with a thin lawn of OP50 bacteria. Animals were allowed to adjust to recover from transferring for at least 30 minutes, and then were recorded for 2 minutes on the plate.

After all crawling plates had been recorded, one milliliter of PBS was added to plates, and animals were recorded swimming for 2 minutes. Videos were processed by WormLab (MBF Bioscience) to detect and track both crawling and swimming animals. Animals recorded crawling continuously for 30 seconds or swimming for 20 seconds continuously were kept for analysis.

### Mitochondrial stress assays in C. elegans

The transgene *Phsp-6::gfp* was used to measure mitochondrial stress (Yoneda et al., 2004). Double balanced heterozygous animals with one extra copy were crossed with wild-type (VC2010), or *atp-1*(extra copy) animals to cross out the *hIn1* balancer and to only have one copy of the *tmC5* balancer except for animals with two extra copies (Fig. S7). 8-10 animals 24 hours post L4 stage were placed per well in a 398 well plate with PBS + 1% pluronic acid that had been stored at the experimental temperature (20° C for P316 and S331, or 15° C for R167).

2.5 mg/mL Levamisole in PBS stored at the experimental temperature was added to each well for a final concentration of 1.5 mg/mL at the same time and immediately taken for imaging in the brightfield and GFP fluorescence channel on the Thermofisher CellInsight CX-7 HCS as previously described (Gosai et al., 2010; Li et al., 2014; Long et al., 2014). The same images were run through two separate image thresholds, one where GFP head fluorescence was the only fluorescence counted in *tmC5/+* animals (labeled as WT in graphs), and a second where GFP head fluorescence + fluorescence in the intestine of control animals was detected.

Average head fluorescence per worm was determined from WT animals. Average head fluorescence for one WT animal was multiplied by number of animals in each well and subtracted from total fluorescence to obtain only the stress fluorescence. Total stress fluorescence was normalized by area of animals to correct for differences in animal size, and then normalized to WT fluorescence to control for differences between replicates.

### Mitochondrial morphology in C. elegans muscle

Larval stage L4 animals homozygous for *bcIs80* and *tmC5* balancer, and heterozygous for the *hIn1* balancer were imaged with a 63x oil objective lens at 3x magnification on 2% agarose pads in 10mM levamisole. Muscles from the dorsal two quadrants were imaged approximately halfway between the pharynx and the vulva. Animals were imaged within 15 minutes of being placed in levamisole. Maximum projections of 4-6 μm z-stacks were used. Z-projections were smoothed once in ImageJ and then processed by the ImageJ MitoSegNet PreProcessing plug in to set image depth. Images were then processed by MitoS CPU for windows program with a 20 pixel minimum size to create image masks, and then analyzed for mitochondrial size using MitoA analyzer and compared using Prism (Fischer et al., 2020). 20-23 images were used for each genotype.

### Mitochondrial assays in fibroblasts

Respiratory chain enzyme activities for complexes I, II, III, II+III, IV and citrate synthase were assayed spectrophotometrically at 30° C in cellular homogenates as previously described and compared to over 30 control fibroblasts (Chatfield et al., 2015; Coughlin et al., 2015).

Complex V activity was determined by measuring the oligomycin-sensitive ATP synthase hydrolytic activity as detailed previously (Mayr et al., 2004; Rustin et al., 1994). In this assay, ATP is converted to ADP and P_i_ by ATP synthase that is coupled via pyruvate kinase and lactate dehydrogenase to the oxidation of NADH to NAD^+^. The oxidation of NADH was followed at 340 nm for 5 min at 37°C, after which oligomycin was added and the assay was followed for an additional 5 min to determine the oligomycin-insensitive ATPase activity.

Solubilized inner mitochondrial membrane fractions were isolated by differential centrifugation from skin fibroblasts and were separated by blue native polyacrylamide gel electrophoresis (BN-PAGE) and then evaluated by in-gel activity staining assays for complexes I, II, IV, and V as described previously (Chatfield et al., 2015; Coughlin et al., 2015; Smet et al., 2009; Van Coster et al., 2001). The assembly of complex I by non-denaturing BN-PAGE followed by western blot analysis using the antibody against NDUFS2 was also analyzed as described (Friederich et al., 2017).

Steady state amounts of the proteins of subunits of complex V catalytic core and peripheral stalk including ATP5PO, the linker between the F_1_ catalytic core and the peripheral stalk, ATP5F1A and ATP5F1B, the α and the β subunits of the F_1_ catalytic core respectively. were assessed by SDS-PAGE followed by western blot as previously described (Ganapathi et al., 2022).

The maximal catalytic ATP synthesis rate was measured based on the method described with some modifications (Rustin et al., 1994). Fibroblasts were permeabilized with digitonin (2 μg/mL for 1 minute), and incubated with 1 mM substrates malate and pyruvate, and 0.2 mM DDAP. The reaction was started by addition of excess ADP 1 mM at 37°C with sampling at time points 0, 1 min, 2 min, 3 min, 5 min, and 15 min. Samples were then deproteinized with PCA, pH neutralized, and ATP was measured following derivatization to ethenoadenosine using bromoacetaldehyde (Sharon et al., 2006), and a Adenine nucleotides were separated using reverse phase HPLC while monitoring fluorescence at λ_exc_ 280nm λ_em_ 410nm. Bromoacetaldehyde was made by refluxing of bromoacetaldehyde diethylacetal in acetic acid environment at 100 °C for 2 hours followed by neutralization to pH 4.3. Adenosine containing analytes were derivatized to ethenoadenosine by combining with 2x volume of the bromoacetaldehyde solution (∼245 mM) followed by incubation at 80 °C for 20 minutes, then cooled and filtered through 0.22 μm centrifugal membranes before HPLC on an Agilent 1100 series. The HPLC separation method involved: 10 μL injection sample volume, 0.7 mL/min flow rate; Atlantis® T3 3.0 x 100 mm column with 3 μM bead size (Waters) controlled at 30 °C. The gradient involved solvent A: 30 mM potassium phosphate pH 5.45 with 0.8 mM tetrabutylammonium phosphate (TBAP, an ion pairing reagent); and solvent B: 1:1 volume by volume of acetonitrile : 30 mM potassium phosphate pH7 with 0.8 mM TBAP in water. After optimization, the following gradient was used: 90/10% A/B from 0-0.1 min, change to 70/30% from 0.1 to 0.7 min, hold at 70/30% from 0.7 to 1.7 min, change to 55/45% from 1.7 to 2.6 min, hold at 55/45% from 2.6 to 5 min, before returning back from 5 to 6 min to initial 90/10% and re-equilibrating for 20 min. Analytes were detected using an Agilent 1200 series fluorescence detector with excitation λ 280 nm and emission λ 410 nm. All adenosine species was resolved with retention times: ADO ∼3.2 min; AMP ∼4.1 min; ADP ∼5.8 min; ATP ∼6.7 min, and two alternative internal standards: 2-methyl-adenosine ∼3.5 min, and 5-acyl-adenosine ∼4.0 min. Using 5 repeat measurements of a standard curve run on 3 separate days, the limit of detection was 500 nM and the limit of quantification was 1 μM for all analytes, and the coefficient of variation for repeat analyses was <1%. The appearance rate of ATP production was used with subtraction of the oligomycin resistant rate.

### Blue native PAGE in lymphoblastoid cell lines

Blue Native PAGE (BN-PAGE) analysis was performed by solubilizing whole cell lymphoblastoid pellets from the proband 2 and two unrelated individual pediatric controls in a digitonin solution containing solubilization buffer (20 mM Bis-Tris (pH 7.0), 50 mM NaCl, 10% glycerol, 1% digitonin (w/v)). Protein concentration was determined using the Pierce Protein Assay Kit (Thermo Fisher Scientific), and a total of 30 ug of whole cell lysate was separated on an Invitrogen NativePAGE Bis-Tris Gel (3-12%), followed by transferring proteins onto PVDF membrane (Merck) using the Invitrogen Mini Blot Module transfer system, as per the manufacturer’s instructions. The gel loading was visualized by staining the PVDF membrane in a stain containing 0.1% Coomassie Brilliant Blue G-250 (CBB; Acros Organics). Immunoblots were developed using primary antibodies for ATP5F1A (Abcam; ab14748) and SDHA (Abcam; ab14715) at 1:1000 dilutions and secondary horseradish peroxidase coupled mouse antibody (Cell Signalling Technology). Images were visualized on a ChemiDoc XRS+ imaging machine (BioRad) using Clarity Western ECL Substrate (BioRad).

### Seahorse analysis of proband fibroblasts

The MitoStress test was conducted on a Seahorse 96XF bioanalyzer according to the manufacturer’s instruction. After performing optimization assays on wild-type fibroblasts for cell number (3,000, 10,000, 15,000 and 20,000 cells) and trifluoromethoxyphenylhydrazone (FCCP) concentration (0.25, 0.5 and 1µM), 15,000 cells per well (seeded 16-24 hours prior to running on the bioanalyzer) and 1µM FCCP were used for all MitoStress Tests comparing fibroblasts from *ATP5F1A* p.R182Q proband and race, sex and age-matched control fibroblasts (GM03349, Coriell Institute for Medical Research).

### TMRE-mitochondrial membrane potential measurements of proband fibroblasts

*ATP5F1A* p.R182Q proband or control fibroblast cells were cultured in DMEM medium supplemented with 15% heat-inactivated FBS (complete medium). Cells were seeded at a density of 1 x 10^4^ cells/well of a 96-well plate. Following an overnight incubation, mitochondrial membrane potential was measured using the TMRE-Mitochondrial membrane potential assay kit (Abcam AB11385) following the manufacturer’s protocol. Briefly, culture medium was removed and replaced with either complete medium or 20µM FCCP for 10 minutes. TMRE at a final concentration of 20nM was added to cells and incubated for 30 mins. Medium was then replaced with 100µl of PBS/0.2% BSA + 5µg/ml Hoechst and incubated for 20 mins before imaging. TMRE fluorescence (peak emission 575nM) was detected by the CX-7 high-content imager (Thermofisher). The cell number was determined by counting the number of nuclei using the Hoechst mask. TMRE fluorescence was normalized to the cell number. All data was further normalized to the untreated wild-type control.

### Cellular ATP measurements of proband fibroblasts

Total cellular ATP was measured using the Luminescent ATP Detection Assay Kit (Abcam, AB113849) according to the manufacturer’s instructions. Briefly, *ATP5F1A* p.R182Q proband or control fibroblast cells were cultured in 96-well plates at a density of 1.4 x 10^4^ cells/well. The next day, detergent (provided in the kit) was added to lyse the cells and stabilized the ATP. Following a 5-min incubation, the substrate solution was added to each well. Plates were wrapped in aluminum foil and further incubated for 10 minutes before quantification. Luminescence was measured on the TECAN Infinite M Plex.

### Acquisition and Analysis of Mass Spectrometry Data

Proteomics analysis in fibroblasts was performed using the patient sample in triplicate and five in-assay controls as described in detail previously (Van Hove et al., 2024). Briefly, after washing fibroblasts in PBS after harvesting, were resuspended in SDS-TEAB buffer, and protein measured (Pierce BCA Protein Assay Kit). Twenty-five µg protein was reduced and alkylated using tris(2-carboxyethyl)phosphine (TCEP) and 2-chloroacetamide, transferred to an S-Trap micro column (Protifi, LLC), and overnight trypsinized. The peptides are eluted from the S-Trap micro column using three elution buffers, dried and resuspended in 2% acetonitrile 0.1% trifluoracetic acid and 3 µL per sample was loaded on an OrbiTrap Eclipse Mass Spectrometer (Thermo Fisher Scientific) for liquid chromatography–tandem mass spectrometry (LC-MS/MS) analysis. After loading onto an Acclaim Pepmap 100 C18 75 µm × 2 cm trapping column, they were chromatographically resolved on-line using an EASY-Spray Pepmap RSLC C18 75 µm × 25 cm 2µm analytical column (Thermo Scientific) using an Ultimate 3000 RSLCnano LC system. Data were acquired in data-independent acquisition (DIA) experiments, full MS resolutions were set to 120,000 at m/z with data acquired in centroid mode using positive polarity in 50 windows with resolution set to 30,000. Raw data were processed using the Spectronaut platform (version 19.6.250122.62635, Sagan, Biognosis) and searched against the UniProt human database (canonical peptides + isoforms, reviewed, 42,447 entries), excluding single hit proteins. For proband 2, lymphoblastoid cells from proband (in triplicate) and four unrelated controls were prepared for proteomics as previously described (Hock et al., 2024). MS2 Quantity from protein groups were imported into Perseus (version 2.0.10.0) and two valid values were filtered for patient and in-assay controls. Log_2_ transformed data was generated in Perseus using MitoCarta3.0 (Rath et al., 2021) + entries and normalization via subtract row cluster means prior to a two-sample t-test using p-value = 0.05 for truncation. Log2 fold-changes were applied to the topographical heatmap using a custom build script (Stroud et al., 2016) against the bovine complex V structure downloaded from the protein data bank PDB: 7AJD (Berman et al., 2000; Spikes et al., 2021). The relative complex abundance (RCA) was calculated and plotted for each complex of the respiratory chain, the small and the large subunit of the mitoribosome and the pyruvate dehydrogenase complex in R (version 4.3.0) and RStudio (version 2023.03.1+446) using a custom build program (Hock et al., 2024). The ATP5F1A range plot was calculated in R using MS2 quantities from ATP5F1A normalized to the mean of mitochondrial proteins annotated with MitoCarta3.0 and relative to the median value of controls as previously described (Hock et al., 2024). To determine the variation in normal controls, the RCA was developed from 23 deidentified normal control fibroblast cells with verified normal standard cellular energetics assays.

### Statistics

Statistics were performed in GraphPad Prism. Data sets were tested for normality, then ANOVA (for normal data), or Kruskal-Wallis (for data sets with at least one non-normal genotype) followed by post-hoc Holm-Sidak testing between genotypes were performed.

## Data Availability

All data produced in the present study are available upon reasonable request to the authors

## ACKNOWLEDGEMENTS

The authors would like to thank the subjects and their families for participation in the study. The authors would also like to thank Mike Nonet for helpful consultation in using RMCE to make the *atp-*1[extra copy] line. Some strains were provided by the CGC, which is funded by NIH Office of Research Infrastructure Programs (P40 OD010440). Research reported in this manuscript was also supported by the NIH Common Fund, through the Office of Strategic Coordination/Office of the NIH Director under award number U54 NS108251 (TS and LSK), from the National Institute of Neurological Disorders and Stroke of the National Institutes of Health U01HG007709 and U01HG007942. This project was supported in part by the Center for Rare and Undiagnosed Genetic Diseases, Children’s Discovery Institute, St Louis Children’s Hospital Foundation (GAS and SCP), NIH NIGMS R01 GM100756 (TS), the Clinical Translational Core (CTC) of the Baylor College of Medicine (BCM) Intellectual and Developmental Disabilities Research Center (IDDRC). The BCM IDDRC is supported by P50 HD103555 from the Eunice Kennedy Shriver National Institute of Child Health and Human Development (NICHD). This study was also funded in part by the Indiana University Grand Challenge Precision Health Initiative. In Colorado, the study was supported by philanthropic support from the Children’s Hospital Colorado Summits for Samantha, and the University of Colorado Foundation, and by a grant from the National Institutes of Health, NIH U54NS078059 for the North American Mitochondrial Disease Consortium (NAMDC) to J.L.K.V.H. NAMDC is part of Rare Diseases Clinical Research Network (RDCRN), an initiative of the Office of Rare Diseases Research (ORDR), NCATS. This consortium is funded through collaboration with NCATS. The work performed in the lab of R.R. is supported by NIH/NCCR 1 S10 OD028538- 01A1 to Nichole Reisdorph. In Melbourne, the study was supported by an Australian National Health and Medical Research Council (NHMRC) Investigator Fellowship (2009732 to DAS) along with funding by Medical Research Future Fund Genomics Health Futures Mission (2016030 to DAS and DHH). We thank the Mito Foundation for the provision of instrumentation through research equipment grants to DAS and DHH and the Melbourne Mass Spectrometry and Proteomics Facility (MMSPF) for the provision of instrumentation and training. Analysis was supported by the Centre for Population Genomics (Garvan Institute of Medical Research and Murdoch Children’s Research Institute) and was funded in part by a National Health and Medical Research Council investigator grant (2009982). The Rare Disease Flagship acknowledges financial support from the Royal Children’s Hospital Foundation [2019-1198 and 2023-1484], the Murdoch Children’s Research Institute, Paula Fox, The Andrew and Geraldine Buxton Foundation and The Pierce Armstrong Foundation. The research conducted at the Murdoch Children’s Research Institute was supported by the Victorian Government’s Operational Infrastructure Support Program.

## APPENDIX SUPPLEMENTARY INFORMATION

### Clinical report

Proband 1 is a male with dystonia, global developmental delay since birth, and mild- moderate intellectual disability. He was diagnosed with cerebral palsy and walks with the aid of braces. His first words were delayed. His examination is significant for dystonia, spasticity, gait apraxia, fluency disorder, poor fine and gross motor skills, limited attention, poor muscle tone, and brisk reflexes with normal sensation, but he is nondysmorphic. He has had a normal EEG, normal brain/spine magnetic resonance imaging (MRI), and normal EMG/NCV studies. Blood lactic acid levels have been mildly elevated; CSF lactate was normal. GDF15 levels were normal. Family history was non-contributory. Clinical duo exome sequencing with the parent identified a heterozygous missense variant in *ATP5F1A* (NM_004046.6:c.545G>A p.(Arg182Gln)). Subsequent targeted maternal studies confirmed the variant was *de novo*. Trio genome sequencing did not identify a second *ATP5F1A* variant, and mitochondrial DNA sequencing was normal.

Proband 2 is a male with moderate to severe global developmental delay. He was born at term via forceps after an uncomplicated pregnancy and was in good condition at birth. Early history included significant central hypotonia with delayed crawling and pulling to stand. He can walk with support, but independent ambulation has not been achieved. He has reduced tone with brisk reflexes in upper and lower limbs and upgoing plantar responses. He has a tented upper lip with hypotonic facies and profound central hypotonia. He displays prominent drooling but there have been no concerns regarding swallowing or possible aspiration. Verbal communication is limited to a handful of words, however comprehension appears better than expressive language. There has been no regression nor seizures. Growth is in the normal range. Serum lactate was borderline at 2.2 mM (NR 0.5-1.4 mM). Genetic testing including chromosome microarray, fragile X testing, trio exome sequencing and *DMPK* repeat analysis was non diagnostic. Trio genome sequencing identified a recurrent *de novo* heterozygous missense variant in *ATP5F1A* (NM_004046.6:c.545G>A p.(Arg182Gln)). Mitochondrial genome sequencing was non diagnostic.

Proband 3 is a female with a significant history of severe global developmental delay and abnormal muscle tone, characterized by central hypotonia and peripheral spasticity. Her medical history is further complicated by dysphagia, chronic constipation, frequent vomiting, and feeding difficulties necessitating the placement of a gastrostomy tube (G-tube). She had a mildly elevated lactate 3.1 mM (normal 0.5-2.2), which at other times were normal. Genetic testing, including chromosomal microarray analysis (CMA) and mitochondrial DNA analysis were unrevealing. Subsequent clinical exome sequencing identified a heterozygous variant of uncertain significance (NM_004046.6:c.1037C>T p.(Ser346Phe)) in the *ATP5F1A* gene. It also identified a single heterozygous pathogenic variant in the *MMAA* (NM_172250.3:c.433C>T p.(Arg145Ter)) gene, but biochemical testing had been normal and did not support the autosomal recessive methylmalonic aciduria diagnosis. Subsequent testing confirmed that the *ATP5F1A* variant in this patient is *de novo*.

Proband 4 is a female who was born prematurely. Developmental delays were noted. Over the next years, she gradually developed spasticity and dystonia, and could only walk with adaptive devices. On exam, she has dystonia most notable in facial and upper extremity movements. She has severe spasticity, most pronounced in her lower extremities, and has contractures in elbows and knees. Her functionality improved following dorsal rhizotomy. She has fatiguability. She is fed by gastrostomy tube for dysphagia and has severe gastroesophageal reflux. At age 11-15 years, she has growth retardation with height at -4 SD and weight at -3.3 SD. She is very socially interactive using assistive devices. Her lactate and her GDF15 levels were normal. She had a normal oxidized/reduced glutathione ratio. However, while non-fasting, she has elevated ketones with 3-hydroxybutyrate 1.96 mM, and acetoacetate 0.50 mM, with a clear elevated ratio of 3.89 (normal 0.47-2.57). Genetic testing showed a de novo variant in *ATP5F1A* (NM_004046.6:c.1037C>T p.(Ser346Phe)).

Proband 5 is a female with significant intellectual disability, developmental regression, growth failure, and history of hospitalizations for ketoacidosis. Prenatally, she was noted to have a dilated right ventricle in her brain, thought to be due to an ischemic event. Neonatally, she had hyperbilirubinemia, which improved with phototherapy. She had failure to thrive, multiple epistaxis which improved with surgery and cautery, an atrial septal defect, sinus tachycardia with normal QTc, mild scoliosis, and she is not dysmorphic. She had one possible seizure event between 6-10 years of age. Developmental concerns were raised early in life. She walked with stiffness and toe walking, which has improved. She is currently nonverbal. She had poor social interaction at around 1-5 years of age, but has improved. Behaviorally, she shows stereotypic hand movements and teeth grinding, and she had abusive behaviors in the past. Treatment with fluoxetine has improved anxiety and crying. On neurologic exam she has asymmetric increased tone on the left side, 3+ reflex on the left patella, ankle clonus on the left, and abnormal gait.

She has had 8 hospitalizations with ketoacidosis (3-hydroxybutyrate 3.6 mM), typically with normal ammonia, with the first episode with PICU admission. Her ketones normalize when she is healthy. She also has intermittent elevations of blood (1.1-5.5 mM) with normal lactate:pyruvate ratio, and urine lactate without a clinical correlate. She has never had documented hypoglycemia but has been hyperglycemic. Brain MRI has shown stable asymmetry of lateral ventricles and stable focal volume loss. GDF15 levels were normal. Skin fibroblast studies for beta ketothiolase, succinyl CoA:3-oxoacid CoA transferase (SCOT), and pyruvate carboxylase (PC) were normal. Clinical trio exome sequencing for proband 5 identified a *de novo* missense variant in *ATP5F1A* (NM_004046.6:c.992C>T p.(Pro331Leu)). Additionally, a *de novo FLT1* consensus splice variant (NM_002019.4:c.1276+2T>C) was identified; this was considered as a potential candidate for her prenatal stroke, given the role of FLT1 (also known as VEGFR1) in vascular development (Chappell et al., 2013; Nesmith et al., 2017). Compound heterozygous variants in *IARS1* were also identified (paternally inherited NM_002161.6:c.2500G>A p.(Val834Met) and maternally inherited NM_002161.6:c.3713C>T p.(Thr1238Ile)). Pathogenic variants in *IARS1* cause growth retardation, impaired intellectual development, hypotonia, and hepatopathy (OMIM #617093). However, zinc deficiency is a common feature of this condition, and the proband had normal zinc levels, and the p.Thr1238Ile variant has been classified as likely benign in ClinVar (variation 744089), so this was not prioritized as a possible contributor (Kopajtich et al., 2016). Finally, a maternally inherited *PCK2* variant (NM_004563.4:c.1405C>T p.(Arg469Cys)) was identified. This was considered as a candidate contributory factor for her ketoacidosis, since recessive *PCK2* variants cause mitochondrial phosphoenolpyruvate carboxykinase deficiency (OMIM #261650); however, a second variant in this gene was not identified. Mitochondrial DNA testing identified one rare homoplasmic *CYTB* variant, m.15099T>C, which has been classified as likely benign in ClinVar (variation 693824), and thus ruled out as contributory.

Proband 6 is a male with global developmental delay, limited verbal communication, postnatal failure to thrive with feeding difficulties, axial hypotonia, and leg spasticity with contractures (and therefore is nonambulatory). Echocardiogram and brain MRI were normal. Metabolic workup, including lactate, ammonia, acylcarnitine profile, and plasma amino acids, was largely unremarkable. Targeted genetic testing, including chromosomal microarray analysis, spinal muscular atrophy testing, ataxia panel, Fragile X testing, and Prader-Willi methylation studies were nondiagnostic. Mitochondrial genome sequencing revealed a 22% heteroplasmic variant of uncertain significance in the gene MT-TI (m.4317A>G), inherited from his parent who has 9% heteroplasmy in blood; this variant is not considered contributory to his phenotype. Trio whole exome sequencing and later whole genome sequencing identified a *de novo* heterozygous missense variant in *ATP5F1A* (NM_004046.6:c.326T>C p.(Leu109Ser)).

## Supplementary Figure Legends

**Figure S1.**
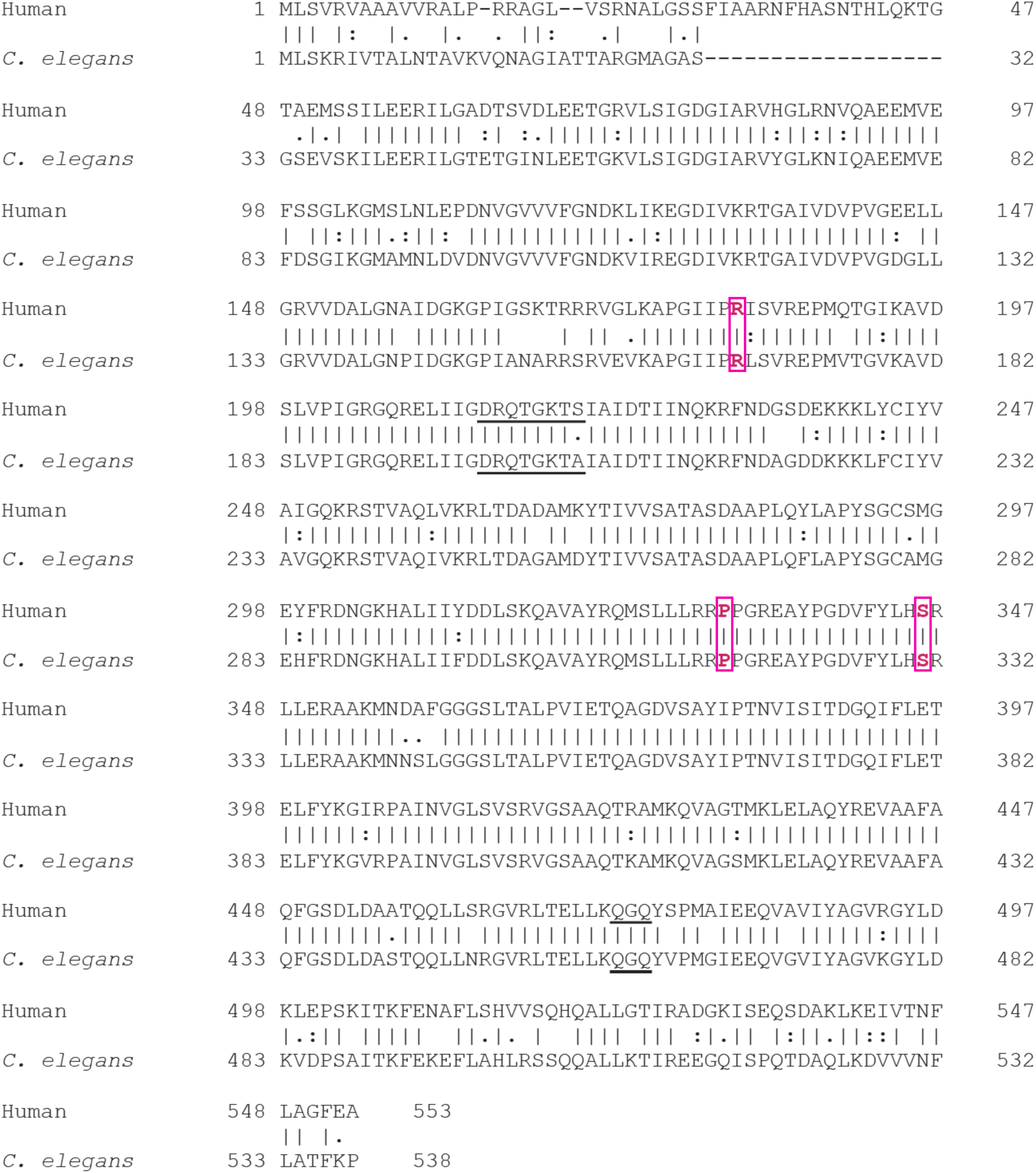
– *ATP5F1A* is highly conserved as *atp-1*. Sequence alignment of human ATP5F1A and *C. elegans* ATP-1 protein sequences. Horizontal dashes indicate gap, vertical dashes indicate identity, colon indicates conservative change, blank indicates mismatch. Wild type proband variant residues are highlighted in magenta. Annotated ATP binding sites from UniProt are underlined.

**Figure S2.**
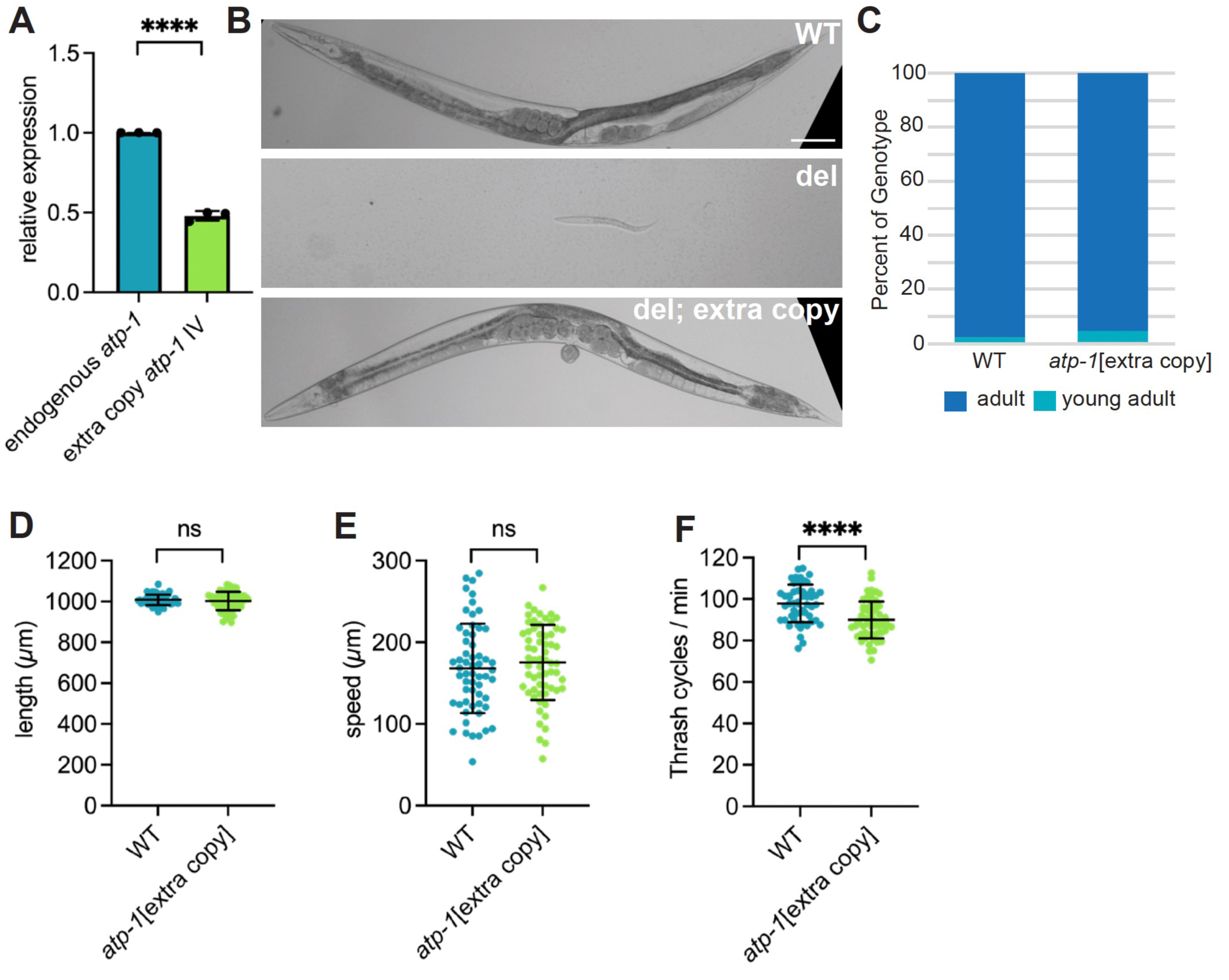
– Animals with extra copies of wild type *atp-1* are superficially normal. (A) Relative expression of endogenous *atp-1* compared to WT *atp-1* transgene inserted on chromosome IV as measured by RNA-seq. The transgene has two synonymous changes compared to the endogenous *atp-1*. (B) Animals 72 hours after embryo laying. WT animals grow to adults at 72 hours post embryo lay (top), while *atp-1* deletion homozygous animals arrest at larval stage L1 (middle). Growth and fertility of *atp-1* deletion animals are fully rescued by addition of two transgenic copies of *atp-1* (*atp-1[extra copy]*) (bottom). Scale bar 100 µm. Quantification of development rate (C), body length (D), crawl speed (E) and thrashing rate of wild type animals with and without extra copies of *atp-1*. ns- not significant, * p <0.05, ** p<0.01, *** p<0.001, **** p < 0.0001.

**Figure S3.**
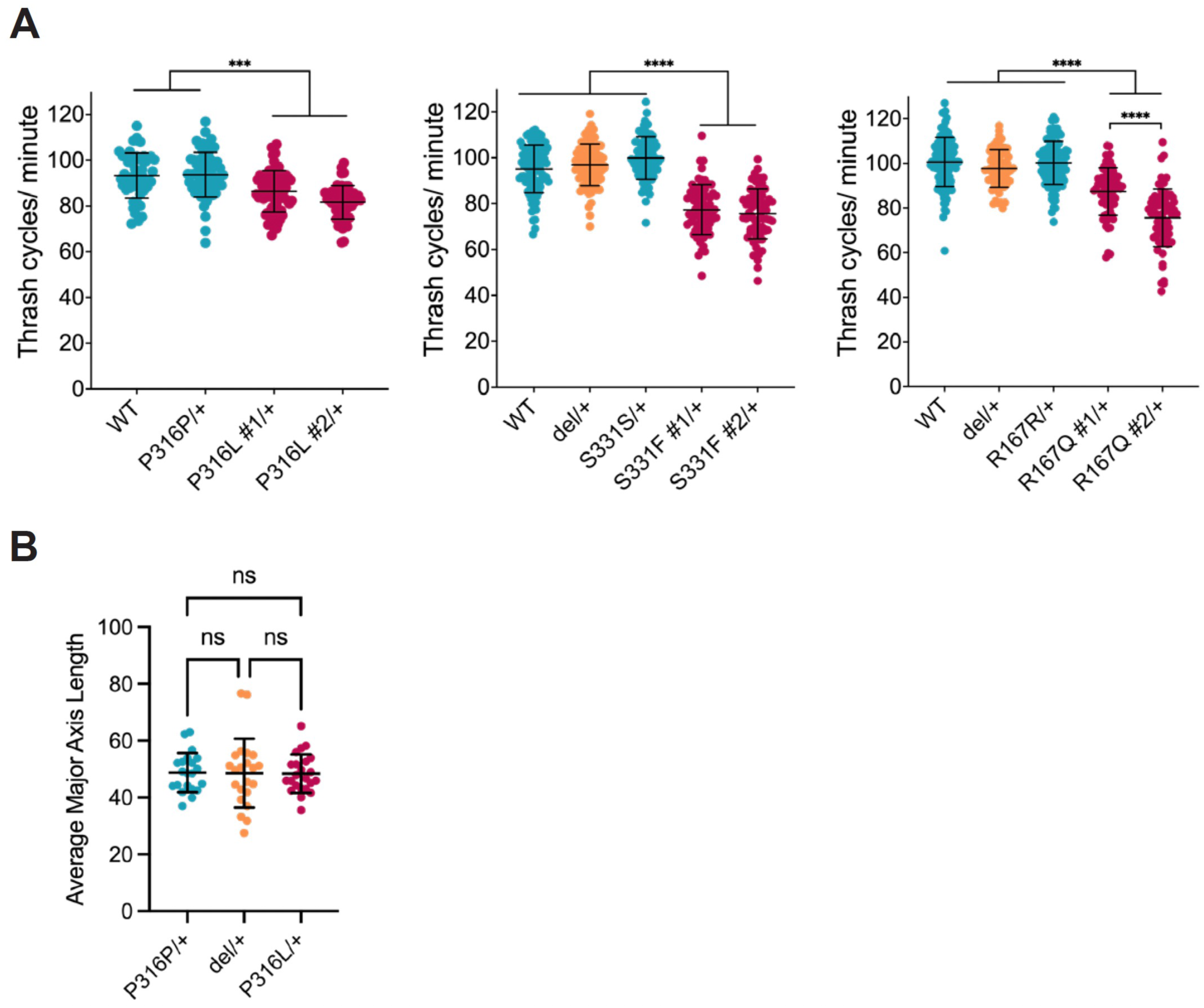
*atp-1* proband variant animals thrash slower and P316L animals have normal mitochondrial tubule length. (A) One day adult proband variant animals allowed to swim in PBS show slower thrashing as compared to control animals. (B) Quantification of major axis length of *C. elegans* muscle mitochondria shows no difference between heterozygous control edit, heterozygous deletion, and heterozygous P316L animals. ns- not significant, * p <0.05, ** p<0.01, *** p<0.001, **** p < 0.0001.

**Figure S4.**
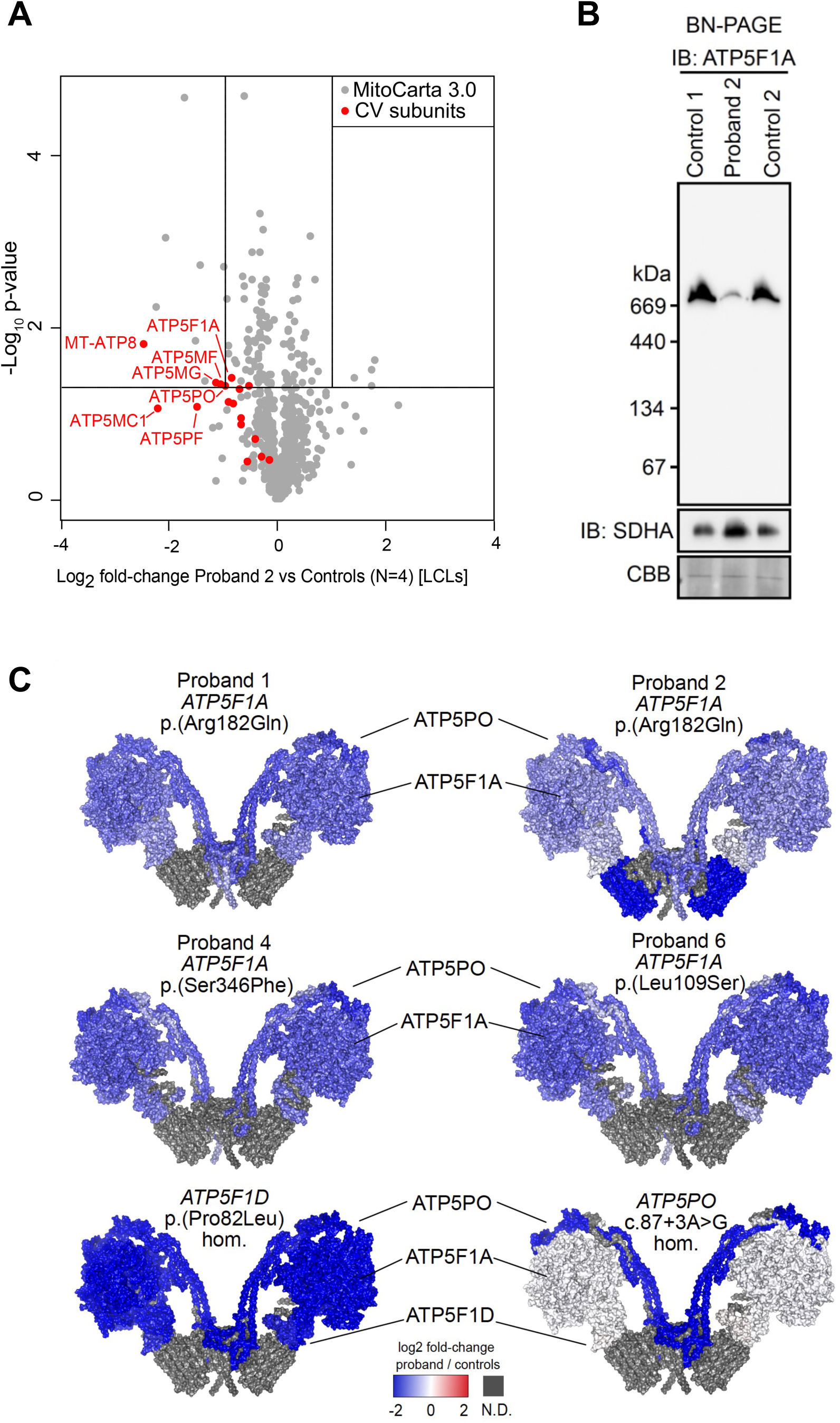
(A) Volcano plot of mitochondrial proteins annotated from MitoCarta3.0 of Proband 2 (p.R182Q) lymphoblastoid cell lines (LCLs) compared to controls (N=4) showing reduced abundance of subunits of Complex V. Vertical lines represent ± 2-fold-change equivalent and horizontal lines represent significance p-value = 0.05 equivalent. Red = Complex V subunits. (B) Blue native PAGE and immunoblotting (IB) of LCLs from Proband 2 and two unrelated controls against ATP5F1A and SDHA antibodies showing reduced abundance of complex V in Proband 2. CBB: Coomassie Brilliant Blue. (C) Topographical heatmap of the log2 fold-change abundances onto the cryo-EM structure of the dimer complex V structure for probands 1, 2, 4 and 6 as well as disease controls with known biallelic pathogenic variants in *ATP5F1D* and *ATP5PO* relative to controls.

**Figure S5.**
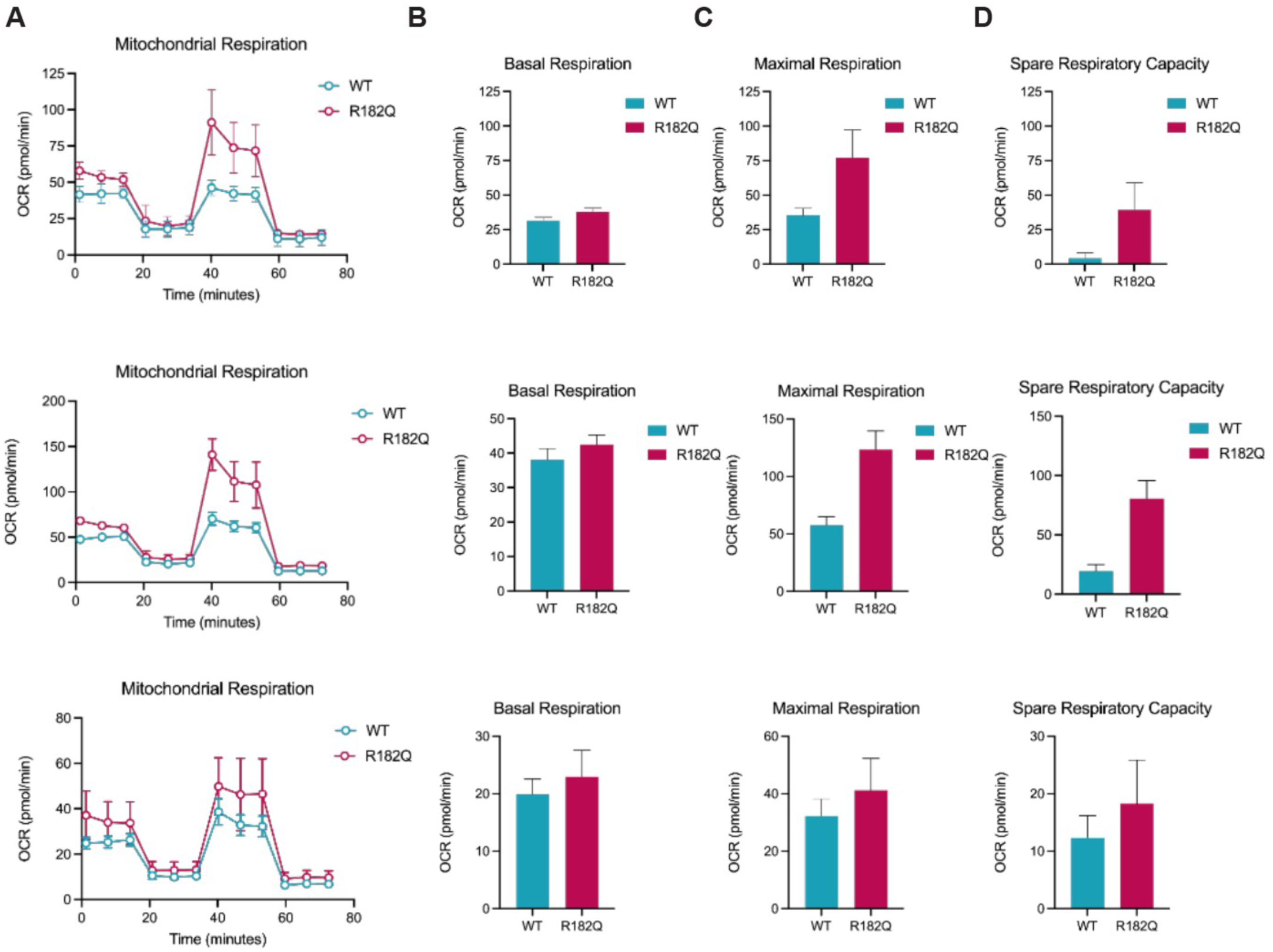
Individual replicates of mitochondrial stress tests performed on proband derived fibroblasts shown in. Fig. 6. (A) Three individual replicates of mitochondrial respiration studies of *ATP5F1A* p.R182Q proband fibroblasts and age, race, and sex matched control fibroblasts as measured by Seahorse. Corresponding (B) Basal, (C) maximal, and (D) spare oxygen consumption rates (OCR) calculated from each experiment in A.

**Figure S6.**
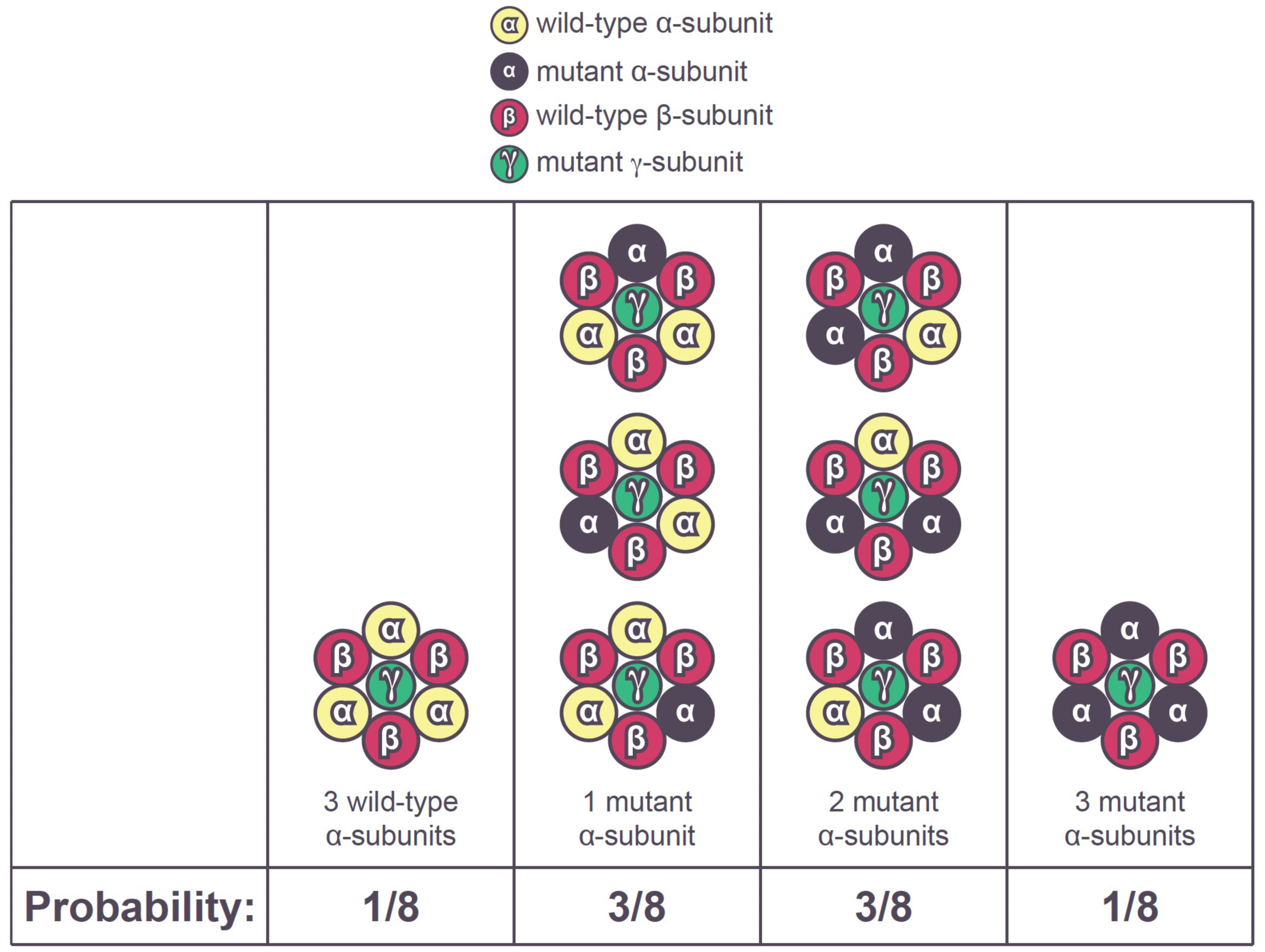
Possible combinations of α- and β-subunit assembly in F_1_ complex. The α-and β-subunits form hetero-hexameric ring around the central stalk (γ-subunit). 1/8^th^ of the complex formed will have 3 wild-type α-subunits. 3/8^th^ will have 1 mutant and two wild-type α-subunits. 3/8^th^ will have 2 mutant and 1 wild-type α-subunits. 1/8^th^ will have 3 mutant α-subunits.

**Figure S7.**
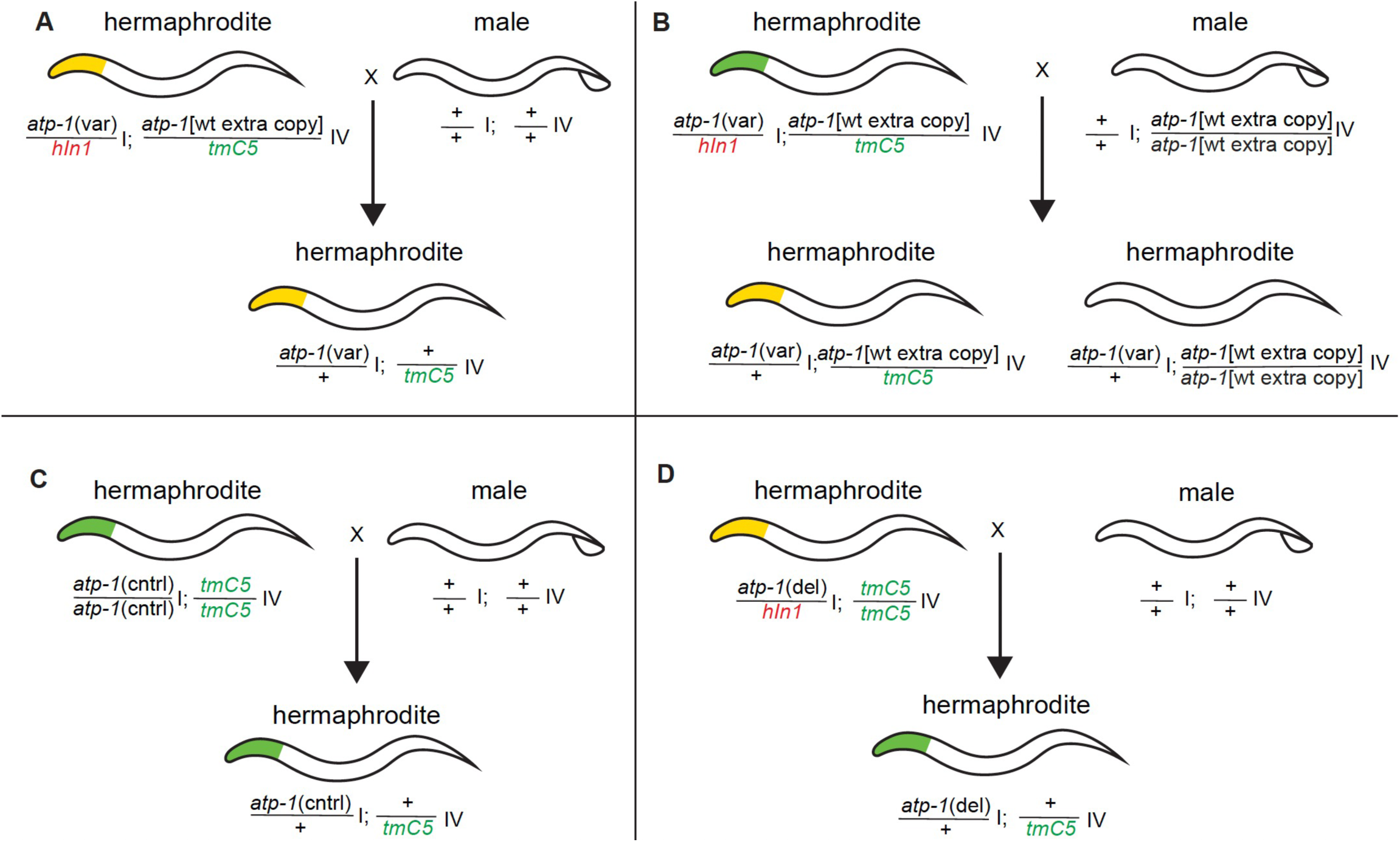
Crossing schematic to generate animals analyzed in this study. Animals heterozygous for the *hIn1* balancer have red heads (pharynx), animals heterozygous for *tmC5* balancer have green heads (pharynx). + indicates locus with no transgene. Animals homozygous for either balancer are uncoordinated. (A) heterozygous proband variant animals with one extra copy that had red and green heads (depicted as yellow) were crossed with WT males to obtain heterozygous animals with no extra copies that were used in Figs. 3, 4, and S3. (B) Heterozygous proband variant animals with one extra copy that had red and green heads were crossed with extra copy males to obtain heterozygous animals with one extra copy and heterozygous animals with two extra copies that were used in Fig. 3. (C) Animals homozygous for the control edits or WT and for the *tmC5* balancer were mated to WT males to obtain control edit animals and WT animals in Figs. 3, 4, and S3. (D) Animals heterozygous for *atp-1* deletion and homozygous for the *tmC5* balancer were crossed with WT males to obtain deletion heterozygotes in Figs. 3, 4, and S3.

**Table S1A.**
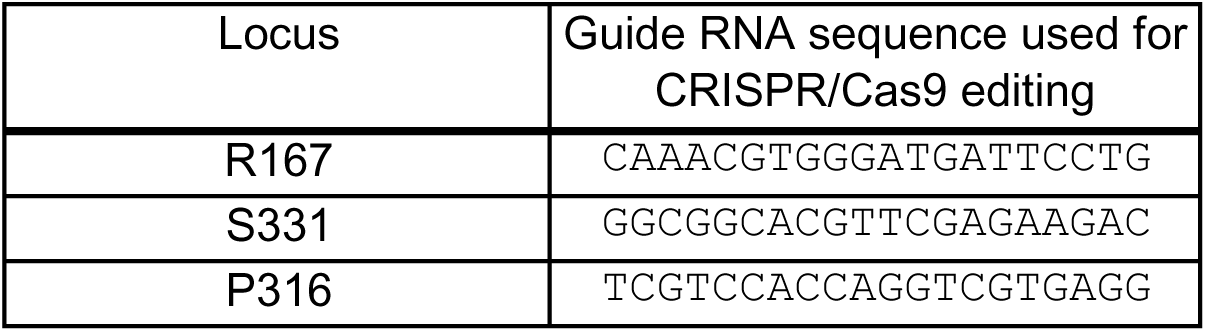
Guide sequences used for CRISPR/Cas9 editing.

**Table S1B.**
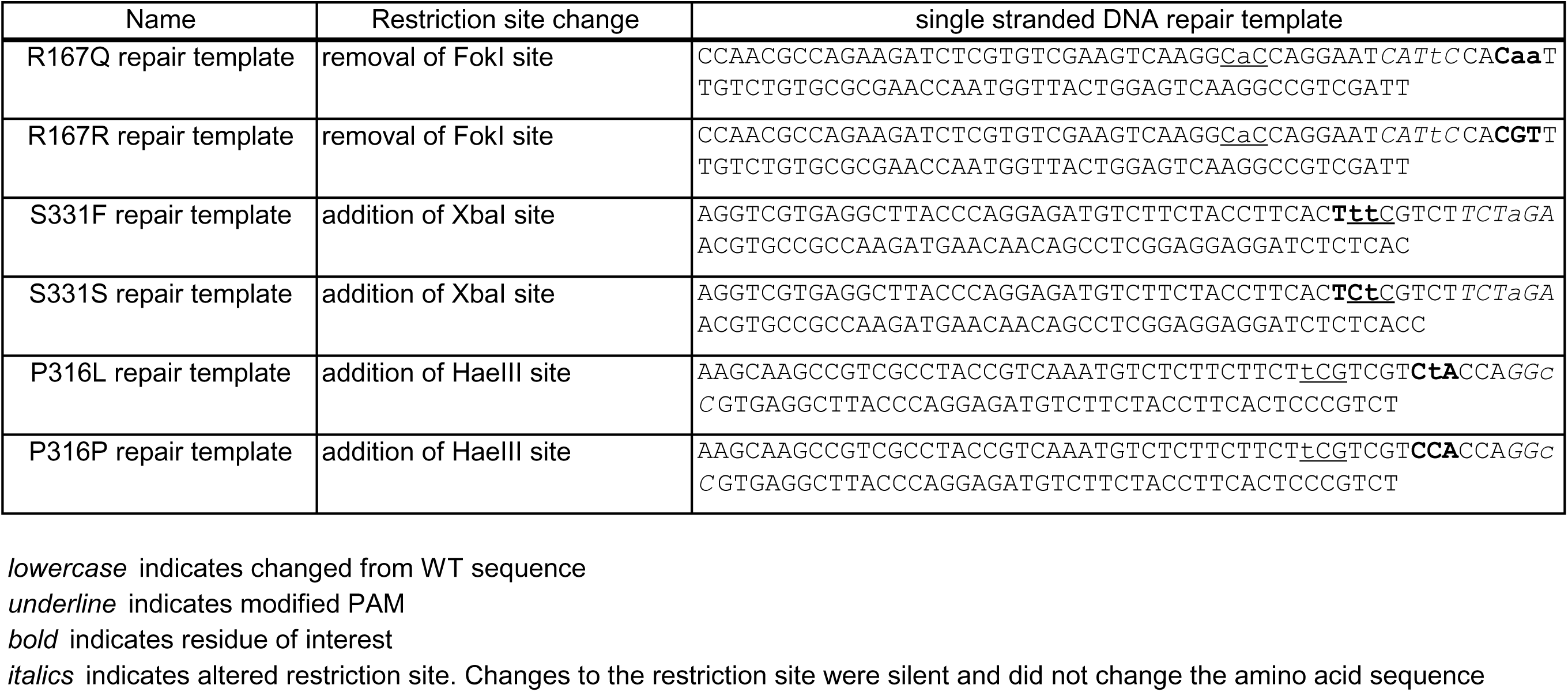
Repair templates used for CRISPR/Cas9 editing.

**Table S2.** Respiratory chain enzyme activities.

| **Proband 1**<br>**p.Arg182Gln** | **Activity (controls)** | **Z-score** | **Ratio /CS** | **Z-score** | **Ratio/complex II** | **Z-score** |
| --- | --- | --- | --- | --- | --- | --- |
| Complex I | 91.0 (49.3-131.1) | 0.2 | 256 (145-396) | 0.5 | 373 (237-754) | -0.1 |
| Complex II | 243.9 (130.9-364.4) | 0.3 | 687 (297-863) | 0.6 | NA | NA |
| Complex III | 17.2 (8-29.2) | 0.5 | 48 (19-65) | 0.7 | 70 (36-114) | 0.4 |
| Complex II-III | 133.1 (61.8-158.8) | 0.8 | 375 (131-376) | 1.2 | 546 (263-1100) | 0.2 |
| Complex IV | 2.1 (2.2-7.1) | -1.9 | 6 (6-23) | -2 | 9 (12-35) | -2.1 |
| Citrate synthase | 355.1 (253.5-554.1) | -0.3 | NA | NA | NA | NA |
| **Complex V hydrolysis** | **131.6 (205.8-440.6) % of avg: 42%** | **-2.76** |  |  |  |  |

| **Proband 4**<br>**p.Ser346Phe** | **Activity (controls)** | **Z-score** | **Ratio /CS** | **Z-score** | **Ratio/complex II** | **Z-score** |
| --- | --- | --- | --- | --- | --- | --- |
| Complex I | 114.1 (49.3-131.1) | 1 | 241 (145-396) | 0.5 | 425 (237-754) | 0.3 |
| Complex II | 268.1 (130.9-364.4) | 0.6 | 566 (297-863) | 0 | NA | NA |
| Complex III | 11.3 (8-29.2) | -0.5 | 24 (19-65) | -0.7 | 42 (36-114) | -0.7 |
| Complex II-III | 13.2 (61.8-158.8) | 0.8 | 286 (131-376) | 0.6 | 504 (263-1100) | 0.1 |
| Complex IV | 5.6 (2.2-7.1) | 0.7 | 12 (6-23) | 0.6 | 21 (12-35) | 0.6 |
| Citrate synthase | 473.4 (253.5-554.1) | 0.7 | NA | NA | NA | NA |
| **Complex V hydrolysis** | **104.6 (205.8-440.6) % of avg: 33%** | **-3.17** |  |  |  |  |

| **Proband 6**<br>**p.Leu109Ser** | **Activity (controls)** | **Z-score** | **Ratio /CS** | **Z-score** | **Ratio/complex II** | **Z-score** |
| --- | --- | --- | --- | --- | --- | --- |
| Complex I | 155.7 (49.3-131.1) | 2 | 467 (145-396) | 2.7 | 725 (237-754) | 1.9 |
| Complex II | 214.3 (130.9-364.4) | -0.1 | 643 (297-863) | 0.3 | NA | NA |
| Complex III | 18.8 (8-29.2) | 0.7 | 56 (19-65) | 1 | 87 (36-114) | 0.8 |
| Complex II-III | 150.8 (61.8-158.8) | 1.2 | 453 (131-376) | 1.8 | 704 (263-1100) | 0.7 |
| Complex IV | 3.6 (2.2-7.1) | -0.5 | 11 (6-23) | -0.4 | 17 (12-35) | -0.5 |
| Citrate synthase | 333.3 (253.5-554.1) | -0.6 | NA | NA | NA | NA |
| **Complex V hydrolysis** | **116.7 (205.8-440.6) % of avg: 37%** | **-2.98** |  |  |  |  |

**Table S3.**
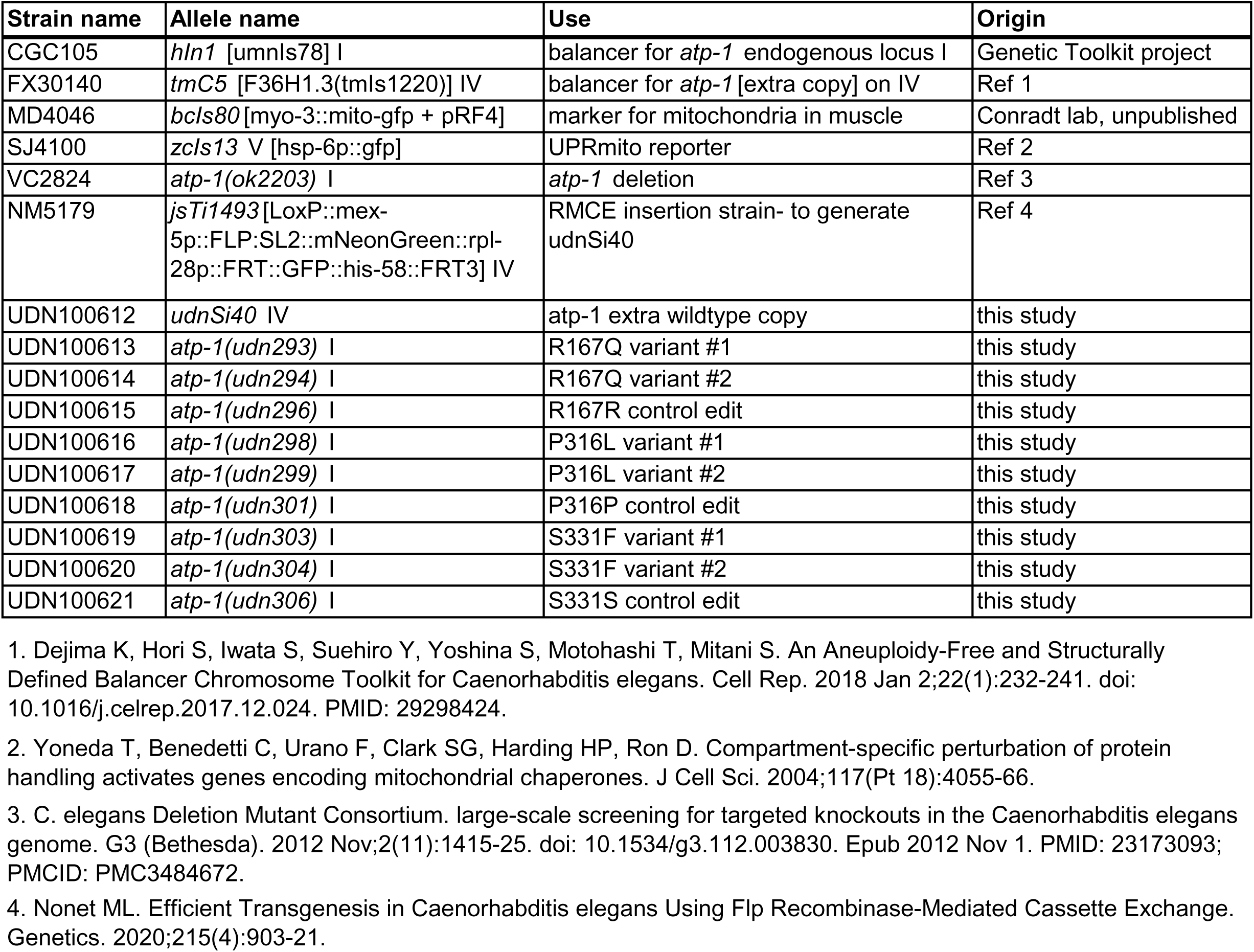
*C. elegans* lines used in study.

**Table S4.**
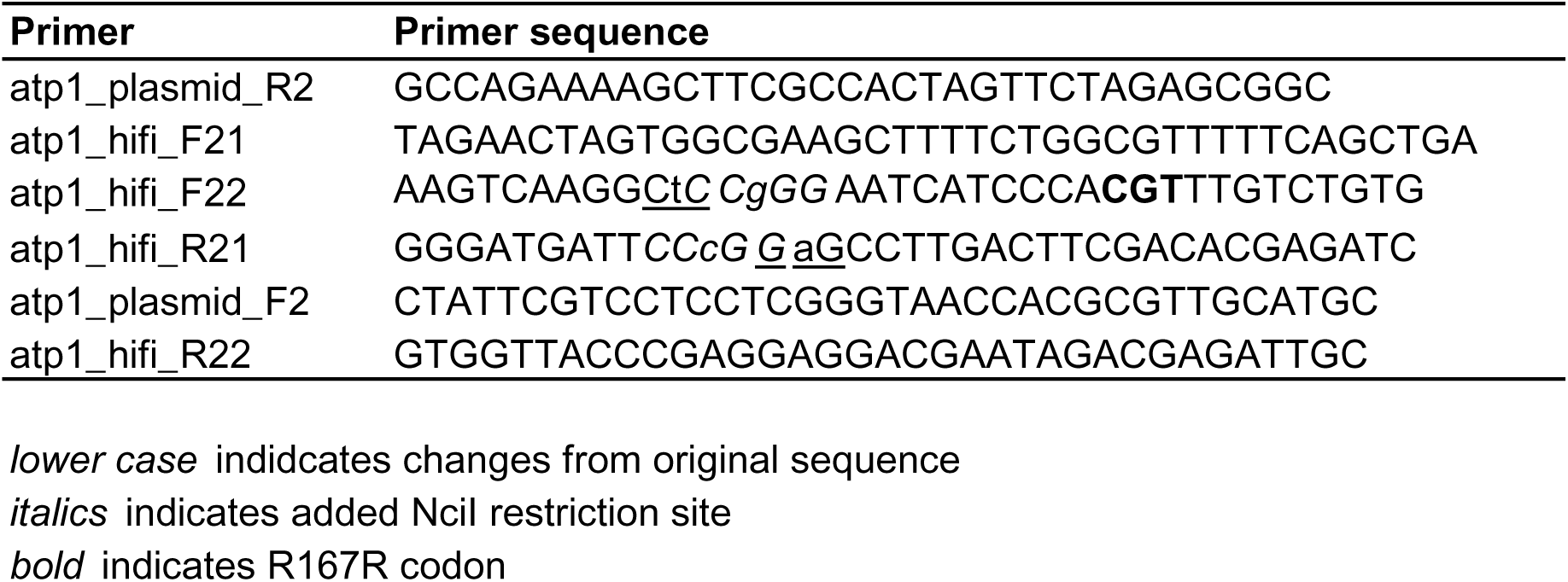
Primers used to clone wild type *atp-1* extra copy.

## BIBLIOGRAPHY

Abrahams, J. P., Leslie, A. G., Lutter, R., & Walker, J. E. (1994). Structure at 2.8 A resolution of F1- ATPase from bovine heart mitochondria. Nature, 370(6491), 621–628. 10.1038/370621a0

Anders, K. R., & Botstein, D. (2001). Dominant-lethal alpha-tubulin mutants defective in microtubule depolymerization in yeast. Mol Biol Cell, 12(12), 3973–3986. 10.1091/mbc.12.12.3973

Anderson, N. S., & Haynes, C. M. (2020). Folding the Mitochondrial UPR into the Integrated Stress Response. Trends Cell Biol, 30(6), 428–439. 10.1016/j.tcb.2020.03.001

Bejsovec, A., & Anderson, P. (1988). Myosin heavy-chain mutations that disrupt Caenorhabditis elegans thick filament assembly. Genes Dev, 2(10), 1307–1317. 10.1101/gad.2.10.1307

Berman, H. M., Westbrook, J., Feng, Z., Gilliland, G., Bhat, T. N., Weissig, H., Shindyalov, I. N., & Bourne, P. E. (2000). The Protein Data Bank. Nucleic Acids Res, 28(1), 235–242. 10.1093/nar/28.1.235

Booth, K. T., Ghaffar, A., Rashid, M., Hovey, L. T., Hussain, M., Frees, K., Renkes, E. M., Nishimura, C. J., Shahzad, M., Smith, R. J., Ahmed, Z., Azaiez, H., & Riazuddin, S. (2020). Novel loss-of-function mutations in COCH cause autosomal recessive nonsyndromic hearing loss. Hum Genet, 139(12), 1565–1574. 10.1007/s00439-020-02197-5

Booth, K. T. A., Schulte, R. R., Smith, L., Gao, H., Stohler, R. A., Liu, Y., Reshmi, S. C., & Vance, G. H. (2024). ZMIZ1::ABL1 Fusion: An Uncommon Molecular Event With Clinical Implications in Pediatric Cancer. Arch Pathol Lab Med. 10.5858/arpa.2024-0082-OA

Boyer, P. D. (1997). The ATP synthase--a splendid molecular machine. Annu Rev Biochem, 66, 717–749. 10.1146/annurev.biochem.66.1.717

Campbell, D., & Zuryn, S. (2024). The mechanisms and roles of mitochondrial dynamics in C. elegans. Semin Cell Dev Biol, 156, 266–275. 10.1016/j.semcdb.2023.10.006

Chatfield, K. C., Coughlin, C. R., 2nd, Friederich, M. W., Gallagher, R. C., Hesselberth, J. R., Lovell, M. A., Ofman, R., Swanson, M. A., Thomas, J. A., Wanders, R. J., Wartchow, E. P., & Van Hove, J. L. (2015). Mitochondrial energy failure in HSD10 disease is due to defective mtDNA transcript processing. Mitochondrion, 21, 1–10. 10.1016/j.mito.2014.12.005

Chen, S., Francioli, L. C., Goodrich, J. K., Collins, R. L., Kanai, M., Wang, Q., Alfoldi, J., Watts, N. A., Vittal, C., Gauthier, L. D., Poterba, T., Wilson, M. W., Tarasova, Y., Phu, W., Grant, R., Yohannes, M. T., Koenig, Z., Farjoun, Y., Banks, E., . . . Karczewski, K. J. (2024). A genomic mutational constraint map using variation in 76,156 human genomes. Nature, 625(7993), 92–100. 10.1038/s41586-023-06045-0

Chen, X. J., & Clark-Walker, G. D. (1995). Specific mutations in alpha- and gamma-subunits of F1- ATPase affect mitochondrial genome integrity in the petite-negative yeast Kluyveromyces lactis. EMBO J, 14(13), 3277–3286. 10.1002/j.1460-2075.1995.tb07331.x

Chen, X. J., & Clark-Walker, G. D. (1996). The mitochondrial genome integrity gene, MGI1, of Kluyveromyces lactis encodes the beta-subunit of F1-ATPase. Genetics, 144(4), 1445–1454. 10.1093/genetics/144.4.1445

Clark-Walker, G. D., Hansbro, P. M., Gibson, F., & Chen, X. J. (2000). Mutant residues suppressing rho(0)-lethality in Kluyveromyces lactis occur at contact sites between subunits of F(1)- ATPase. Biochim Biophys Acta, 1478(1), 125–137. 10.1016/s0167-4838(00)00003-0

Coughlin, C. R., 2nd, Scharer, G. H., Friederich, M. W., Yu, H. C., Geiger, E. A., Creadon-Swindell, G., Collins, A. E., Vanlander, A. V., Coster, R. V., Powell, C. A., Swanson, M. A., Minczuk, M., Van Hove, J. L., & Shaikh, T. H. (2015). Mutations in the mitochondrial cysteinyl-tRNA synthase gene, CARS2, lead to a severe epileptic encephalopathy and complex movement disorder. J Med Genet, 52(8), 532–540. 10.1136/jmedgenet-2015-103049

Dautant, A., Meier, T., Hahn, A., Tribouillard-Tanvier, D., di Rago, J. P., & Kucharczyk, R. (2018). ATP Synthase Diseases of Mitochondrial Genetic Origin. Front Physiol, 9, 329. 10.3389/fphys.2018.00329

Fielder, S. M., Rosenfeld, J. A., Burrage, L. C., Emrick, L., Lalani, S., Attali, R., Bembenek, J. N., Hoang, H., Baldridge, D., Silverman, G. A., Undiagnosed Diseases, N., Schedl, T., & Pak, S. C. (2022). Functional analysis of a novel de novo variant in PPP5C associated with microcephaly, seizures, and developmental delay. Mol Genet Metab, 136(1), 65–73. 10.1016/j.ymgme.2022.03.007

Fischer, C. A., Besora-Casals, L., Rolland, S. G., Haeussler, S., Singh, K., Duchen, M., Conradt, B., & Marr, C. (2020). MitoSegNet: Easy-to-use Deep Learning Segmentation for Analyzing Mitochondrial Morphology. iScience, 23(10), 101601. 10.1016/j.isci.2020.101601

Friederich, M. W., Erdogan, A. J., Coughlin, C. R., 2nd, Elos, M. T., Jiang, H., O’Rourke, C. P., Lovell, M. A., Wartchow, E., Gowan, K., Chatfield, K. C., Chick, W. S., Spector, E. B., Van Hove, J. L. K., & Riemer, J. (2017). Mutations in the accessory subunit NDUFB10 result in isolated complex I deficiency and illustrate the critical role of intermembrane space import for complex I holoenzyme assembly. Hum Mol Genet, 26(4), 702–716. 10.1093/hmg/ddw431

Galber, C., Carissimi, S., Baracca, A., & Giorgio, V. (2021). The ATP Synthase Deficiency in Human Diseases. Life (Basel), 11(4). 10.3390/life11040325

Ganapathi, M., Friocourt, G., Gueguen, N., Friederich, M. W., Le Gac, G., Okur, V., Loaec, N., Ludwig, T., Ka, C., Tanji, K., Marcorelles, P., Theodorou, E., Lignelli-Dipple, A., Voisset, C., Walker, M. A., Briere, L. C., Bourhis, A., Blondel, M., LeDuc, C., . . . Chung, W. K. (2022). A homozygous splice variant in ATP5PO, disrupts mitochondrial complex V function and causes Leigh syndrome in two unrelated families. J Inherit Metab Dis, 45(5), 996–1012. 10.1002/jimd.12526

Ganetzky, R. D., Markhard, A. L., Yee, I., Clever, S., Cahill, A., Shah, H., Grabarek, Z., To, T. L., & Mootha, V. K. (2022). Congenital Hypermetabolism and Uncoupled Oxidative Phosphorylation. N Engl J Med, 387(15), 1395–1403. 10.1056/NEJMoa2202949

Gomes, L. C., Di Benedetto, G., & Scorrano, L. (2011). During autophagy mitochondria elongate, are spared from degradation and sustain cell viability. Nat Cell Biol, 13(5), 589–598. 10.1038/ncb2220

Gosai, S. J., Kwak, J. H., Luke, C. J., Long, O. S., King, D. E., Kovatch, K. J., Johnston, P. A., Shun, T. Y., Lazo, J. S., Perlmutter, D. H., Silverman, G. A., & Pak, S. C. (2010). Automated high-content live animal drug screening using C. elegans expressing the aggregation prone serpin alpha1-antitrypsin Z. PLoS One, 5(11), e15460. 10.1371/journal.pone.0015460

Haynes, C. M., Yang, Y., Blais, S. P., Neubert, T. A., & Ron, D. (2010). The matrix peptide exporter HAF-1 signals a mitochondrial UPR by activating the transcription factor ZC376.7 in C. elegans. Mol Cell, 37(4), 529–540. 10.1016/j.molcel.2010.01.015

Hock, D. H., Caruana, N. J., Semcesen, L. N., Lake, N. J., Formosa, L. E., Amarasekera, S. S. C., Stait, T., Tregoning, S., Frajman, L. E., Robinson, D. R. L., Ball, M., Reljic, B., Ryder, B., Wallis, M. J., Vasudevan, A., Beck, C., Peters, H., Lee, J., Tan, N. B., . . . Stroud, D. A. (2024). Untargeted proteomics enables ultra-rapid variant prioritization in mitochondrial and other rare diseases. medRxiv, 2024.2008.2006.24311318. 10.1101/2024.08.06.24311318

Honzik, T., Tesarova, M., Mayr, J. A., Hansikova, H., Jesina, P., Bodamer, O., Koch, J., Magner, M., Freisinger, P., Huemer, M., Kostkova, O., van Coster, R., Kmoch, S., Houstek, J., Sperl, W., & Zeman, J. (2010). Mitochondrial encephalocardio-myopathy with early neonatal onset due to TMEM70 mutation. Arch Dis Child, 95(4), 296–301. 10.1136/adc.2009.168096

Huang, H., Pan, J., Spielberg, D. R., Hanchard, N. A., Scott, D. A., Burrage, L. C., Dai, H., Murdock, D., Rosenfeld, J. A., Mohammad, A., Huang, T., Lindsey, A. G., Kim, H., Chen, J., Ramu, A., Morrison, S. A., Dawson, Z. D., Hu, A. Z., Tycksen, E., . . . Schedl, T. (2022). A dominant negative variant of RAB5B disrupts maturation of surfactant protein B and surfactant protein C. Proc Natl Acad Sci U S A, 119(6). 10.1073/pnas.2105228119

Ioannidis, N. M., Rothstein, J. H., Pejaver, V., Middha, S., McDonnell, S. K., Baheti, S., Musolf, A., Li, Q., Holzinger, E., Karyadi, D., Cannon-Albright, L. A., Teerlink, C. C., Stanford, J. L., Isaacs, W. B., Xu, J., Cooney, K. A., Lange, E. M., Schleutker, J., Carpten, J. D., . . . Sieh, W. (2016). REVEL: An Ensemble Method for Predicting the Pathogenicity of Rare Missense Variants. Am J Hum Genet, 99(4), 877–885. 10.1016/j.ajhg.2016.08.016

Jacobs, A., Burns, C., Patel, P., Treat, K., Helm, B. M., Conboy, E., & Vetrini, F. (2022). Reanalysis of a novel variant in the IGF1R gene in a family with variable prenatal and postnatal growth retardation and dysmorphic features: benefits and feasibility of IUSM-URDC (Undiagnosed Rare Disease Clinic) program. Cold Spring Harb Mol Case Stud, 8(2). 10.1101/mcs.a006170

Jaganathan, K., Kyriazopoulou Panagiotopoulou, S., McRae, J. F., Darbandi, S. F., Knowles, D., Li, Y. I., Kosmicki, J. A., Arbelaez, J., Cui, W., Schwartz, G. B., Chow, E. D., Kanterakis, E., Gao, H., Kia, A., Batzoglou, S., Sanders, S. J., & Farh, K. K. (2019). Predicting Splicing from Primary Sequence with Deep Learning. Cell, 176(3), 535–548 e524. 10.1016/j.cell.2018.12.015

Jonckheere, A. I., Renkema, G. H., Bras, M., van den Heuvel, L. P., Hoischen, A., Gilissen, C., Nabuurs, S. B., Huynen, M. A., de Vries, M. C., Smeitink, J. A., & Rodenburg, R. J. (2013). A complex V ATP5A1 defect causes fatal neonatal mitochondrial encephalopathy. Brain, 136(Pt 5), 1544–1554. 10.1093/brain/awt086

Keehan, L., Jiang, M. M., Li, X., Marom, R., Dai, H., Murdock, D., Liu, P., Hunter, J. V., Heaney, J. D., Robak, L., Emrick, L., Lotze, T., Blieden, L. S., Undiagnosed Diseases, N., Lewis, R. A., Levin, A. V., Capasso, J., Craigen, W. J., Rosenfeld, J. A., . . . Burrage, L. C. (2021). A novel de novo intronic variant in ITPR1 causes Gillespie syndrome. Am J Med Genet A, 185(8), 2315–2324. 10.1002/ajmg.a.62232

Kemphues, K. J., Raff, E. C., Raff, R. A., & Kaufman, T. C. (1980). Mutation in a testis-specific beta-tubulin in Drosophila: analysis of its effects on meiosis and map location of the gene. Cell, 21(2), 445–451. 10.1016/0092-8674(80)90481-x

Kuhlbrandt, W. (2019). Structure and Mechanisms of F-Type ATP Synthases. Annu Rev Biochem, 88, 515–549. 10.1146/annurev-biochem-013118-110903

Lai, Y., Zhang, Y., Zhou, S., Xu, J., Du, Z., Feng, Z., Yu, L., Zhao, Z., Wang, W., Tang, Y., Yang, X., Guddat, L. W., Liu, F., Gao, Y., Rao, Z., & Gong, H. (2023). Structure of the human ATP synthase. Mol Cell, 83(12), 2137–2147 e2134. 10.1016/j.molcel.2023.04.029

Li, J., Pak, S. C., O’Reilly, L. P., Benson, J. A., Wang, Y., Hidvegi, T., Hale, P., Dippold, C., Ewing, M., Silverman, G. A., & Perlmutter, D. H. (2014). Fluphenazine reduces proteotoxicity in C. elegans and mammalian models of alpha-1-antitrypsin deficiency. PLoS One, 9(1), e87260. 10.1371/journal.pone.0087260

Lieber, D. S., Calvo, S. E., Shanahan, K., Slate, N. G., Liu, S., Hershman, S. G., Gold, N. B., Chapman, B. A., Thorburn, D. R., Berry, G. T., Schmahmann, J. D., Borowsky, M. L., Mueller, D. M., Sims, K. B., & Mootha, V. K. (2013). Targeted exome sequencing of suspected mitochondrial disorders. Neurology, 80(19), 1762–1770. 10.1212/WNL.0b013e3182918c40

Liesa, M., & Shirihai, O. S. (2013). Mitochondrial dynamics in the regulation of nutrient utilization and energy expenditure. Cell Metab, 17(4), 491–506. 10.1016/j.cmet.2013.03.002

Lines, M. A., Cuillerier, A., Chakraborty, P., Naas, T., Duque Lasio, M. L., Michaud, J., Pileggi, C., Harper, M. E., Burelle, Y., Toler, T. L., Sondheimer, N., Crawford, H. P., Millan, F., & Geraghty, M. T. (2021). A recurrent de novo ATP5F1A substitution associated with neonatal complex V deficiency. Eur J Hum Genet, 29(11), 1719–1724. 10.1038/s41431-021-00956-0

Liu, Y. J., McIntyre, R. L., Janssens, G. E., & Houtkooper, R. H. (2020). Mitochondrial fission and fusion: A dynamic role in aging and potential target for age-related disease. Mech Ageing Dev, 186, 111212. 10.1016/j.mad.2020.111212

Long, O. S., Benson, J. A., Kwak, J. H., Luke, C. J., Gosai, S. J., O’Reilly, L. P., Wang, Y., Li, J., Vetica, A. C., Miedel, M. T., Stolz, D. B., Watkins, S. C., Zuchner, S., Perlmutter, D. H., Silverman, G. A., & Pak, S. C. (2014). A C. elegans model of human alpha1-antitrypsin deficiency links components of the RNAi pathway to misfolded protein turnover. Hum Mol Genet, 23(19), 5109–5122. 10.1093/hmg/ddu235

Lorenz, C., Lesimple, P., Bukowiecki, R., Zink, A., Inak, G., Mlody, B., Singh, M., Semtner, M., Mah, N., Aure, K., Leong, M., Zabiegalov, O., Lyras, E. M., Pfiffer, V., Fauler, B., Eichhorst, J., Wiesner, B., Huebner, N., Priller, J., . . . Prigione, A. (2017). Human iPSC-Derived Neural Progenitors Are an Effective Drug Discovery Model for Neurological mtDNA Disorders. Cell Stem Cell, 20(5), 659–674 e659. 10.1016/j.stem.2016.12.013

Lund, A. M., Nicholls, A. C., Schwartz, M., & Skovby, F. (1997). Parental mosaicism and autosomal dominant mutations causing structural abnormalities of collagen I are frequent in families with osteogenesis imperfecta type III/IV. Acta Paediatr, 86(7), 711–718. 10.1111/j.1651-2227.1997.tb08573.x

Magner, M., Dvorakova, V., Tesarova, M., Mazurova, S., Hansikova, H., Zahorec, M., Brennerova, K., Bzduch, V., Spiegel, R., Horovitz, Y., Mandel, H., Eminoglu, F. T., Mayr, J. A., Koch, J., Martinelli, D., Bertini, E., Konstantopoulou, V., Smet, J., Rahman, S., . . . Honzik, T. (2015). TMEM70 deficiency: long-term outcome of 48 patients. J Inherit Metab Dis, 38(3), 417–426. 10.1007/s10545-014-9774-8

Marom, R., Zhang, B., Washington, M. E., Song, I. W., Burrage, L. C., Rossi, V. C., Berrier, A. S., Lindsey, A., Lesinski, J., Nonet, M. L., Chen, J., Baldridge, D., Silverman, G. A., Sutton, V. R., Rosenfeld, J. A., Tran, A. A., Hicks, M. J., Murdock, D. R., Dai, H., . . . Lee, B. H. (2023). Dominant negative variants in KIF5B cause osteogenesis imperfecta via down regulation of mTOR signaling. PLoS Genet, 19(11), e1011005. 10.1371/journal.pgen.1011005

Mayr, J. A., Paul, J., Pecina, P., Kurnik, P., Forster, H., Fotschl, U., Sperl, W., & Houstek, J. (2004). Reduced respiratory control with ADP and changed pattern of respiratory chain enzymes as a result of selective deficiency of the mitochondrial ATP synthase. Pediatr Res, 55(6), 988–994. 10.1203/01.pdr.0000127016.67809.6b

Miyazono, Y., Hirashima, S., Ishihara, N., Kusukawa, J., Nakamura, K. I., & Ohta, K. (2018). Uncoupled mitochondria quickly shorten along their long axis to form indented spheroids, instead of rings, in a fission-independent manner. Sci Rep, 8(1), 350. 10.1038/s41598-017-18582-6

Molina, A. J., Wikstrom, J. D., Stiles, L., Las, G., Mohamed, H., Elorza, A., Walzer, G., Twig, G., Katz, S., Corkey, B. E., & Shirihai, O. S. (2009). Mitochondrial networking protects beta-cells from nutrient-induced apoptosis. Diabetes, 58(10), 2303–2315. 10.2337/db07-1781

Muller, H. J. (1932). Further studies on the nature and causes of gene mutations.

Nasca, A., Mencacci, N. E., Invernizzi, F., Zech, M., Keller Sarmiento, I. J., Legati, A., Frascarelli, C., Bustos, B. I., Romito, L. M., Krainc, D., Winkelmann, J., Carecchio, M., Nardocci, N., Zorzi, G., Prokisch, H., Lubbe, S. J., Garavaglia, B., & Ghezzi, D. (2023). Variants in ATP5F1B are associated with dominantly inherited dystonia. Brain, 146(7), 2730–2738. 10.1093/brain/awad068

Nonet, M. L. (2020). Efficient Transgenesis in Caenorhabditis elegans Using Flp Recombinase-Mediated Cassette Exchange. Genetics, 215(4), 903–921. 10.1534/genetics.120.303388

Olahova, M., Yoon, W. H., Thompson, K., Jangam, S., Fernandez, L., Davidson, J. M., Kyle, J. E., Grove, M. E., Fisk, D. G., Kohler, J. N., Holmes, M., Dries, A. M., Huang, Y., Zhao, C., Contrepois, K., Zappala, Z., Fresard, L., Waggott, D., Zink, E. M., . . . Wheeler, M. T. (2018). Biallelic Mutations in ATP5F1D, which Encodes a Subunit of ATP Synthase, Cause a Metabolic Disorder. Am J Hum Genet, 102(3), 494–504. 10.1016/j.ajhg.2018.01.020

Pais, L. S., Snow, H., Weisburd, B., Zhang, S., Baxter, S. M., DiTroia, S., O’Heir, E., England, E., Chao, K. R., Lemire, G., Osei-Owusu, I., VanNoy, G. E., Wilson, M., Nguyen, K., Arachchi, H., Phu, W., Solomonson, M., Mano, S., O’Leary, M., . . . O’Donnell-Luria, A. (2022). seqr: A web-based analysis and collaboration tool for rare disease genomics. Hum Mutat, 43(6), 698–707. 10.1002/humu.24366

Rambold, A. S., Kostelecky, B., Elia, N., & Lippincott-Schwartz, J. (2011). Tubular network formation protects mitochondria from autophagosomal degradation during nutrient starvation. Proc Natl Acad Sci U S A, 108(25), 10190–10195. 10.1073/pnas.1107402108

Rath, S., Sharma, R., Gupta, R., Ast, T., Chan, C., Durham, T. J., Goodman, R. P., Grabarek, Z., Haas, M. E., Hung, W. H. W., Joshi, P. R., Jourdain, A. A., Kim, S. H., Kotrys, A. V., Lam, S. S., McCoy, J. G., Meisel, J. D., Miranda, M., Panda, A., . . . Mootha, V. K. (2021). MitoCarta3.0: an updated mitochondrial proteome now with sub-organelle localization and pathway annotations. Nucleic Acids Res, 49(D1), D1541–D1547. 10.1093/nar/gkaa1011

Rolland, S. G., Motori, E., Memar, N., Hench, J., Frank, S., Winklhofer, K. F., & Conradt, B. (2013). Impaired complex IV activity in response to loss of LRPPRC function can be compensated by mitochondrial hyperfusion. Proc Natl Acad Sci U S A, 110(32), E2967–2976. 10.1073/pnas.1303872110

Rolland, S. G., Schneid, S., Schwarz, M., Rackles, E., Fischer, C., Haeussler, S., Regmi, S. G., Yeroslaviz, A., Habermann, B., Mokranjac, D., Lambie, E., & Conradt, B. (2019). Compromised Mitochondrial Protein Import Acts as a Signal for UPR(mt). Cell Rep, 28(7), 1659–1669 e1655. 10.1016/j.celrep.2019.07.049

Roussel, N., Sprenger, J., Tappan, S. J., & Glaser, J. R. (2014). Robust tracking and quantification of C. elegans body shape and locomotion through coiling, entanglement, and omega bends. Worm, 3(4), e982437. 10.4161/21624054.2014.982437

Rustin, P., Chretien, D., Bourgeron, T., Gerard, B., Rotig, A., Saudubray, J. M., & Munnich, A. (1994). Biochemical and molecular investigations in respiratory chain deficiencies. Clin Chim Acta, 228(1), 35–51. 10.1016/0009-8981(94)90055-8

Schultz, J., Lee, S. J., Cole, T., Hoang, H. D., Vibbert, J., Cottee, P. A., Miller, M. A., & Han, S. M. (2017). The secreted MSP domain of C. elegans VAPB homolog VPR-1 patterns the adult striated muscle mitochondrial reticulum via SMN-1. Development, 144(12), 2175–2186. 10.1242/dev.152025

Schwarz, J. M., Cooper, D. N., Schuelke, M., & Seelow, D. (2014). MutationTaster2: mutation prediction for the deep-sequencing age. Nat Methods, 11(4), 361–362. 10.1038/nmeth.2890

Sharon, E., Levesque, S. A., Munkonda, M. N., Sevigny, J., Ecke, D., Reiser, G., & Fischer, B. (2006). Fluorescent N2,N3-epsilon-adenine nucleoside and nucleotide probes: synthesis, spectroscopic properties, and biochemical evaluation. Chembiochem, 7(9), 1361–1374. 10.1002/cbic.200600070

Smet, J., Seneca, S., De Paepe, B., Meulemans, A., Verhelst, H., Leroy, J., De Meirleir, L., Lissens, W., & Van Coster, R. (2009). Subcomplexes of mitochondrial complex V reveal mutations in mitochondrial DNA. Electrophoresis, 30(20), 3565–3572. 10.1002/elps.200900213

Spikes, T. E., Montgomery, M. G., & Walker, J. E. (2021). Interface mobility between monomers in dimeric bovine ATP synthase participates in the ultrastructure of inner mitochondrial membranes. Proc Natl Acad Sci U S A, 118(8). 10.1073/pnas.2021012118

Stroud, D. A., Surgenor, E. E., Formosa, L. E., Reljic, B., Frazier, A. E., Dibley, M. G., Osellame, L. D., Stait, T., Beilharz, T. H., Thorburn, D. R., Salim, A., & Ryan, M. T. (2016). Accessory subunits are integral for assembly and function of human mitochondrial complex I. Nature, 538(7623), 123–126. 10.1038/nature19754

Taylor, R. C., Berendzen, K. M., & Dillin, A. (2014). Systemic stress signalling: understanding the cell non-autonomous control of proteostasis. Nat Rev Mol Cell Biol, 15(3), 211–217. 10.1038/nrm3752

Tondera, D., Grandemange, S., Jourdain, A., Karbowski, M., Mattenberger, Y., Herzig, S., Da Cruz, S., Clerc, P., Raschke, I., Merkwirth, C., Ehses, S., Krause, F., Chan, D. C., Alexander, C., Bauer, C., Youle, R., Langer, T., & Martinou, J. C. (2009). SLP-2 is required for stress-induced mitochondrial hyperfusion. EMBO J, 28(11), 1589–1600. 10.1038/emboj.2009.89

Van Coster, R., Smet, J., George, E., De Meirleir, L., Seneca, S., Van Hove, J., Sebire, G., Verhelst, H., De Bleecker, J., Van Vlem, B., Verloo, P., & Leroy, J. (2001). Blue native polyacrylamide gel electrophoresis: a powerful tool in diagnosis of oxidative phosphorylation defects. Pediatr Res, 50(5), 658–665. 10.1203/00006450-200111000-00020

Van Hove, J. L. K., Friederich, M. W., Hock, D. H., Stroud, D. A., Caruana, N. J., Christians, U., Schniedewind, B., Michel, C. R., Reisdorph, R., Lopez Gonzalez, E. D. J., Brenner, C., Donovan, T. E., Lee, J. C., Chatfield, K. C., Larson, A. A., Baker, P. R., 2nd, McCandless, S. E., & Moore Burk, M. F. (2024). ACAD9 treatment with bezafibrate and nicotinamide riboside temporarily stabilizes cardiomyopathy and lactic acidosis. Mitochondrion, 78, 101905. 10.1016/j.mito.2024.101905

Wang, Y., Singh, U., & Mueller, D. M. (2007). Mitochondrial genome integrity mutations uncouple the yeast Saccharomyces cerevisiae ATP synthase. J Biol Chem, 282(11), 8228–8236. 10.1074/jbc.M609635200

Weir, H. J., Yao, P., Huynh, F. K., Escoubas, C. C., Goncalves, R. L., Burkewitz, K., Laboy, R., Hirschey, M. D., & Mair, W. B. (2017). Dietary Restriction and AMPK Increase Lifespan via Mitochondrial Network and Peroxisome Remodeling. Cell Metab, 26(6), 884–896 e885. 10.1016/j.cmet.2017.09.024

Wittig, I., Meyer, B., Heide, H., Steger, M., Bleier, L., Wumaier, Z., Karas, M., & Schagger, H. (2010). Assembly and oligomerization of human ATP synthase lacking mitochondrial subunits a and A6L. Biochim Biophys Acta, 1797(6-7), 1004–1011. 10.1016/j.bbabio.2010.02.021

Yoneda, T., Benedetti, C., Urano, F., Clark, S. G., Harding, H. P., & Ron, D. (2004). Compartment-specific perturbation of protein handling activates genes encoding mitochondrial chaperones. J Cell Sci, 117(Pt 18), 4055–4066. 10.1242/jcs.01275

Yoshida, M., Muneyuki, E., & Hisabori, T. (2001). ATP synthase--a marvellous rotary engine of the cell. Nat Rev Mol Cell Biol, 2(9), 669–677. 10.1038/35089509

Zech, M., Kopajtich, R., Steinbrucker, K., Bris, C., Gueguen, N., Feichtinger, R. G., Achleitner, M. T., Duzkale, N., Perivier, M., Koch, J., Engelhardt, H., Freisinger, P., Wagner, M., Brunet, T., Berutti, R., Smirnov, D., Navaratnarajah, T., Rodenburg, R. J. T., Pais, L. S., . . . Prokisch, H. (2022). Variants in Mitochondrial ATP Synthase Cause Variable Neurologic Phenotypes. Ann Neurol, 91(2), 225–237. 10.1002/ana.26293

## BIBLIOGRAPHY

Chappell, J. C., Mouillesseaux, K. P., & Bautch, V. L. (2013). Flt-1 (vascular endothelial growth factor receptor-1) is essential for the vascular endothelial growth factor-Notch feedback loop during angiogenesis. Arterioscler Thromb Vasc Biol, 33(8), 1952–1959. 10.1161/ATVBAHA.113.301805

Kopajtich, R., Murayama, K., Janecke, A. R., Haack, T. B., Breuer, M., Knisely, A. S., Harting, I., Ohashi, T., Okazaki, Y., Watanabe, D., Tokuzawa, Y., Kotzaeridou, U., Kolker, S., Sauer, S., Carl, M., Straub, S., Entenmann, A., Gizewski, E., Feichtinger, R. G., . . . Staufner, C. (2016). Biallelic IARS Mutations Cause Growth Retardation with Prenatal Onset, Intellectual Disability, Muscular Hypotonia, and Infantile Hepatopathy. Am J Hum Genet, 99(2), 414–422. 10.1016/j.ajhg.2016.05.027

Nesmith, J. E., Chappell, J. C., Cluceru, J. G., & Bautch, V. L. (2017). Blood vessel anastomosis is spatially regulated by Flt1 during angiogenesis. Development, 144(5), 889–896. 10.1242/dev.145672

